# Comprehensive detection of genetic and epigenetic alterations in cancer using long reads with TumorLens

**DOI:** 10.64898/2026.03.18.26348569

**Authors:** Luis F Paulin, Minyi Shi, Yilei Fu, Xinchang Zheng, George Au-Yeung, David Bowtell, Jieming Chen, Yuxin Liang, Christian Hammer, Fritz J Sedlazeck

## Abstract

Accurately resolving the full spectrum of somatic alterations remains a major barrier in cancer genomics. Current short-read sequencing methods often prioritize SNVs and copy-number changes while overlooking SVs, haplotype-specific events, and epigenetic dysregulation. To bridge this gap, we present TumorLens, the first unified long-read framework that jointly detects SNVs, indels, SVs, large CNVs, loss-of-heterozygosity, and CpG methylation in a single assay. TumorLens introduces purity-aware long-read CNV/LoH modeling and personalized HLA-locus reconstruction, enabling the mechanistic interpretation of immune escape through allele-specific methylation profiling. Benchmarked across GIAB standards and clinical cohorts, TumorLens accurately recovered key somatic events, including interferon locus disruptions and HLA loss. Furthermore, it revealed pervasive global hypomethylation alongside focal hypermethylation in critical oncogenic pathways. By consolidating multi-omic layers into an end-to-end analytic pipeline, TumorLens establishes a new standard for comprehensive tumor profiling, accelerating the translation of long-read sequencing into precision oncology.

## Introduction

Cancer is fundamentally driven by genomic instability, manifesting as a broad spectrum of somatic mutations that range from single nucleotide substitutions to large-scale chromosomal rearrangements, accompanied by epigenetic alterations such as DNA methylation changes. These genetic alterations can disrupt key signaling pathways and regulatory networks, driving oncogenesis and tumor progression^1–3^. Concurrently, epigenetic modifications play a pivotal role in shaping the cancer epigenome, influencing gene expression patterns and genomic stability^4^. Yet, most molecular analyses of tumors continue to focus predominantly on single nucleotide variants (SNVs) and copy number variants (CNVs)^5^. This narrow scope overlooks other essential mutation classes, including small insertions/deletions (indels), and structural variants (SVs) ranging from tens of base pairs to hundreds of kilobases. Furthermore, existing studies often ignore DNA methylation changes that can profoundly alter gene regulation, chromatin structure, and antigen presentation. Understanding the intricate interplay between genetic mutations and epigenetic modifications is essential for unraveling the molecular underpinnings of cancer. An integrated approach has the potential to not only elucidate the mechanistic basis of tumorigenesis but also to identify novel therapeutic targets and biomarkers.

Long-read sequencing technologies now enable direct observation of large-scale rearrangements and methylation states on individual DNA molecules, but their application in cancer genomics remains at an early stage ^3^. While previous work has highlighted complex SVs as well as extrachromosomal DNA (ecDNA) and other formations of novel molecules^6–8^, clinical adoption has been limited. This is partly due to the fact that most established somatic variant-calling frameworks, optimized for short-read data, are incompatible with long-read sequencing. Consequently, long-read analytical methods continue to lag behind short-read approaches, which already offer robust, routinely implemented tools for CNV detection, loss of heterozygosity (LoH) analysis, and Human Leukocyte Antigen (HLA) typing. One major factor that has often been ignored is tumor purity when reporting results for long-read based methods. SV callers such as Sniffles^9^, Severus^10^ or SV comparison methods such as Truvari^11^, cannot take tumor purity into account when reporting somatic SV. This is despite the fact that tumor purity can have profound impacts on the accuracy of variant calling and interpretation^12^. As a result, the full potential of long-read sequencing to decode the molecular architecture of cancer genomes has yet to be fully realized.

The Human Leukocyte Antigen (HLA) locus represents one of the most polymorphic and structurally complex regions of the human genome, with allelic variants that underpin immune surveillance and tumor evasion. Yet, interrogating cancer-relevant loci such as the Major Histocompatibility Complex (MHC), which houses these highly polymorphic genes, remains challenging due to extreme allelic diversity, repetitive architecture, and extensive structural variation. Short-read sequencing and conventional variant-calling frameworks are poorly suited to resolve this complexity, including allele-specific methylation and intricate haplotype structures. Despite major advances in long-read sequencing for alignment, variant calling, and phasing, robust solutions for cancer-specific phenomena, such as somatic LoH, copy-number imbalance, and purity-dependent mosaicism remain elusive both within the HLA locus and genome-wide. In addition to the classical HLA molecules, several other genes encode proteins involved in various stages of the antigen processing and presentation pathway, such as beta-2 microglobulin (B2M), transporter associated with antigen processing (TAP), and endoplasmic reticulum aminopeptidase (ERAP)^13^. The coordinated function of these proteins ensures the correct processing, loading, and presentation of antigenic peptides onto HLA molecules for T cell recognition^14^. Alterations in these antigen presentation machinery (APM) components can lead to impaired immune recognition of pathogens or tumor cells, thereby compromising the effectiveness of immune responses^15^.

To address these limitations, we developed TumorLens, an integrated framework for genome-wide tumor characterization from long-read sequencing data. TumorLens jointly analyzes SNVs, indels, structural variants, copy number changes, allelic imbalance, and DNA methylation within the same reference framework, while explicitly modeling tumor purity and accommodating datasets with or without matched normal samples. Furthermore, in order to account for the complexity of antigen presentation in cancer, TumorLens has dedicated functionality to detect somatic HLA loss of heterozygosity (LoH), as well as genetic and epigenetic defects in APM genes. We highlight several novelties of TumorLens and showcase them using tumor-matched and tumor-only tissues, revealing multiple key insights across different cancer types. By providing a unified view of genetic and epigenetic alterations, TumorLens enables comprehensive and quantitative profiling of tumor genomes, revealing both locus-specific insights, such as HLA-related immune escape mechanisms, and genome-wide patterns that shape tumor evolution and heterogeneity.

## Results

### Somatic variant detection in cancer with TumorLens

**TumorLens** is a computational pipeline designed to operate directly on raw Oxford Nanopore (ONT) long reads from either tumor-normal pairs or tumor-only samples, generating high-fidelity variant calls and comprehensive somatic profiles. The pipeline utilizes a series of automated, parallelizable modules (see **Methods**) to aggregate genetic and epigenetic variants for integrated analysis. It introduces novel methodological advancements, most notably the first long-read implementation of somatic CNV and LoH detection, which is enabled by novel adaptations of the Spectre CNV caller that explicitly account for tumor purity. (see **Methods** and **Supplement Text**). TumorLens also introduces genome-wide methylation profiling, integrated with variant detection to reveal synergistic genetic-epigenetic tumor signatures (**Figure 1a**). A central feature of the framework is its focus on the antigen presentation machinery (APM); it analyzes 13 HLA class I and II genes alongside 61 additional APM genes (**Supplementary table 1**). This is further optimized by an approach that reconstructs a personalized reference genome containing sample-specific HLA alleles. This personalized reference addresses the challenges of high allelic diversity, facilitating more accurate haplotype discovery and methylation profiling within the HLA locus (see **Methods**). By bridging genetic and epigenetic signals, TumorLens establishes a unified framework for long-read cancer genomics, providing a foundational approach to investigate the mechanisms of tumorigenesis and potential therapeutic vulnerabilities.

As no existing benchmark encompasses the full range of TumorLens’s features, we evaluated the pipeline’s performance using three distinct cell line cohorts (**Supplementary Table 2**). These included: (1) two well-characterized Genome in a Bottle (GIAB) reference samples, HG002 and HG003 (**Supplementary Text and Supplementary Table 3**); (2) the recently released HG008 pancreatic adenocarcinoma cell line and its matched normal control; and (3) the commercial H2009 lung adenocarcinoma cell line with its corresponding blood-derived normal control (BL2009))^16^.

Recently, the GIAB Consortium released a preliminary benchmark (v.0.4) for HG008, a pancreatic ductal adenocarcinoma cell line isolated from a female patient that contains 50 CNV/LoH events ranging from 210 bases to 190.2Mb. TumorLens only reports CNVs ≥ 100kb, thus, only events meeting this size threshold were included in the CNV benchmark (13 excluded). Our pipeline detected 1,717 somatic SVs, 80 somatic CNVs and 22 tumor LoH regions. Our pipeline successfully detected 28/37 somatic CNVs present in the HG008 CNV benchmark, achieving a recall of 75.7%. Furthermore, detailed inspections revealed specific genomic complexities. When compared to the matched normal genome of the same individual, these included one CNV split as two events, three CNVs were reported as larger events where the breakpoints overlapped with identified LoH regions, and three CNVs having an expected CNV close to diploid (**Supplementary table 4**).

Finally, we sequenced the well-characterized lung adenocarcinoma cell line NCI H2009 (23× coverage) together with its matched blood-derived normal control NCI BL2009 (31× coverage). H2009 is derived from a female patient diagnosed with stage IV lung adenocarcinoma, and is a representative of the high LoH rates frequently observed in Lung cancers ^17^. The TumorLens pipeline successfully detected all 20 previously reported LoH regions with breakpoints within 1kb. This represents a significant improvement in precision over previous reports that relied on microsatellite marker analysis (**Supplementary Table 5**). Additionally, TumorLens identified 32 somatic CNVs and accurately classified the HLA class I genes as homozygous: HLA-A*03:01, HLA-B*07:02 and HLA-C*07:02, a finding consistent with established literature ^18^.

In summary, the TumorLens pipeline demonstrated high accuracy in assessing somatic variants in all three benchmark sample sets. It takes the TumorLens pipeline an average of 6 hours, in a high-performance cluster, from alignment to somatic analysis results for processing 30x ONT tumor/normal paired samples. This indicates that TumorLens provides a rapid solution for long-read cancer genomics without compromising on precision.

### Analysis of patient-derived cell lines and tumor purity assessment

We applied TumorLens to characterize three high grade serous ovarian (HGSC) cancer cell line samples patient-derived cell lines from the Australian Ovarian Cancer Study19 (AOCS): AOCS21, AOCS9, and AOCS19. HGSC are among the most chromosomally unstable solid cancers and therefore a useful resource to evaluate TumorLens performance. We present detailed results for AOCS21 as a representative case (additional details are provided in the **Supplementary Text**). In AOCS21, TumorLens identified a complex somatic landscape comprising 44,929 SNVs, 2,491 SVs, 81 CNVs, and 18 LoH regions. Notably, large-scale CNVs (>10 Mb) were detected across 18 autosomes, involving chromosomal arm losses, whole-chromosome losses, and extensive duplications (**Supplementary Table 6**). These findings were corroborated by multiple chromosome-wide LoH regions, including a significant event on Chromosome 6. Here, TumorLens resolved a duplication affecting both the HLA class I genes (1.3 Mb duplication; CN=4) and the interferon (IFN) gene locus. The integrated HLA typing results confirmed that all HLA genes were homozygous in the tumor (**Supplementary Table 7**). The successful detection and cross-validation of this complex duplication/LoH event supported by coverage analysis, SNV data, and HLA typing underscores the analytical versatility of TumorLens.

A significant novelty of TumorLens is the integration of epigenetics profiling (**Figure 1b**). identifies cancer-specific differentially methylated regions (cDMRs) when the proportion of methylated sites within a given genomic window differs by at least 33 percentage points between the tumor and its matched normal counterpart (see **Methods** for details). In AOCS21, TumorLens reported 26% of the genome as cDMRs. Global and locus-specific hyper- and hypomethylation in tumors are known drivers of oncogene or immune-regulatory gene activation^7^,significantly influencing tumor biology and tumor-immune interactions. As an example, we observed a hypomethylation of the Interferon alpha-7 gene (*IFNA7*) in the AOCS21 tumor sample, suggesting a potential epigenetic activation of this interferon gene. Type I interferon genes, such as *IFN-α*, perform complex, context-dependent roles in ovarian cancer, frequently alternating between anti-tumor immune stimulation and the promotion of tumor progression ^20^.

**Figure 1.**
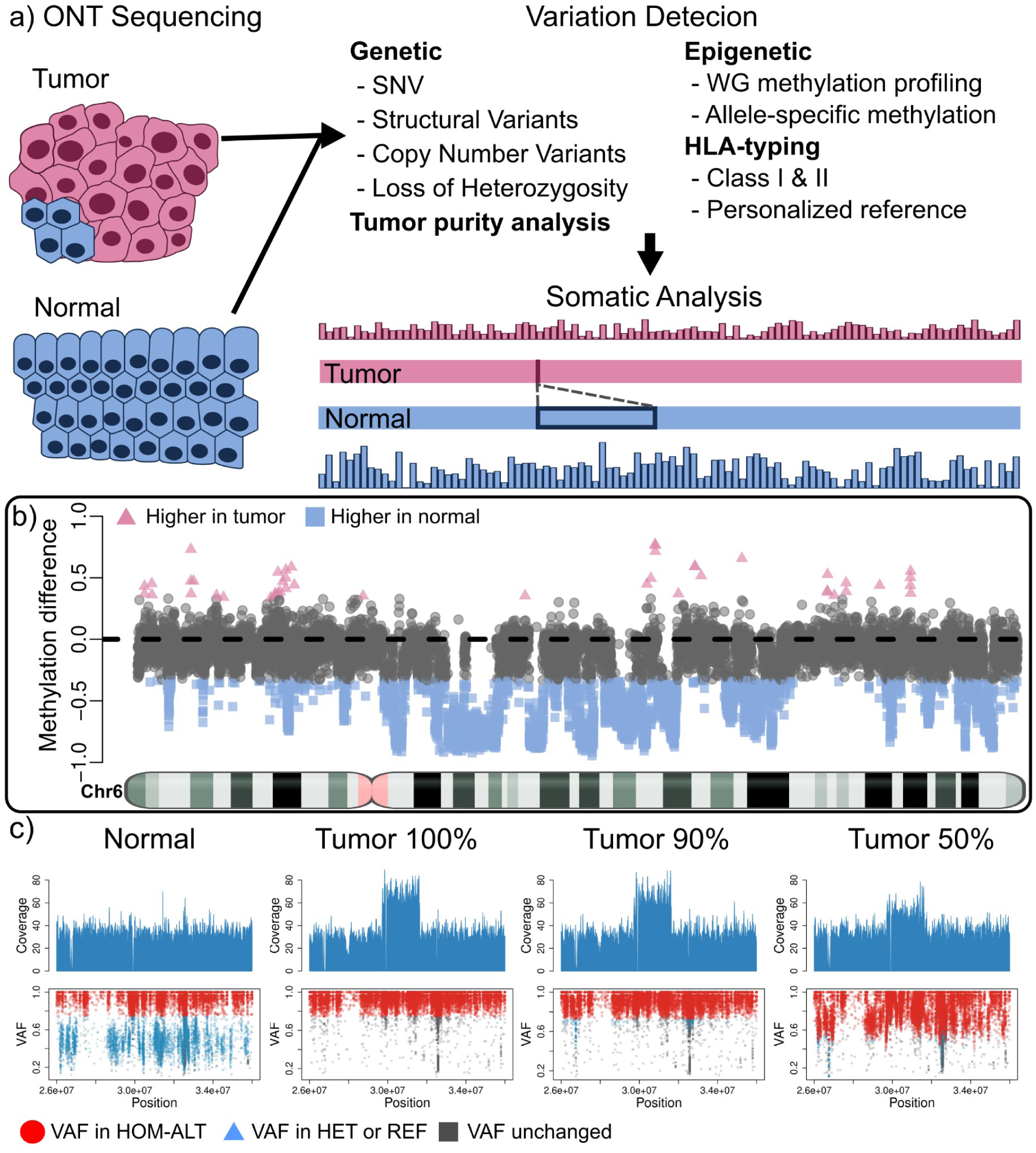
Detection of somatic genetic and epigenetic variations with TumorLens. **A)** TumorLens utilizes ONT sequencing of tumor-normal sample pairs to assess integrated genetic and epigenetic somatic variation in the tumor sample, with further targeted analysis of HLA class I and II genes. Tumor samples can be specified to contain a known proportion of contamination from the normal tissue to improve the analysis. The schematic shows the tumor sample (top, pink) which has lower methylation (barplot) and a detected deletion when compared to the normal tissue (bottom, blue) **B)** Differential methylation assessment shown over Chr 6 of the AOCS21 sample. Methylation differences are shown between tumor and normal for non-overlapping regions. Pink triangles are regions that have higher methylation in the tumor, while blue squares show higher methylation in the normal (gray is undefined). Notice that most of the assessed regions have higher methylation in the normal sample, showing a clear de-regulation of methylation. **C)** Visualization of a duplication (DUP) and Loss of Heterozygosity (LoH) overlapping the HLA gene locus in the ovarian sample AOCS21, compared to the normal and two titration experiments (90%, and 50% tumor purity) to exemplify levels of contamination. The top panel shows the coverage pattern. Notice that the coverage for the DUP is reduced as tumor purity decreases. TumorLens reports a copy number of 4 (CN4) in the 100% tumor sample and a CN3 in the 50% tumor content experiment. The bottom panel shows the SNVs utilized for detecting the LoH traced from the tumor sample to the normal and titration experiments. In red are shown SNVs which have the VAF to be considered homozygous alternative alleles after accounting for tumor content. By incorporating the estimated tumor content, TumorLens can improve the LoH detection even at 50% tumor content. In blue are SNVs that are catalogued as homozygous alternative alleles in the 100% tumor but shown as heterozygous in the normal sample. Gray points represent unchanged heterozygous sites. Acronyms: SNV, single nucleotide variant; WG, whole genome; VAF, variant allele fraction; HOM-ALT, homozygous alternative; HET, heterozygous; REF, reference.

Failure to account for tumor purity has been shown to confound interpretation of both genetic variants and epigenetic alterations^12^. To address this, TumorLens incorporates purity estimates into the variants calling process, by modeling the varying proportion of malignant and non-malignant cells. (see **Methods**). To evaluate pipeline performance across varying levels of cellular heterogeneity, we performed *in silico* titration simulations using ovarian samples AOCS21 and AOCS9, with tumor concentrations ranging from 90% down to 10% (See **Supplementary Table 6**). As illustrated in **Figure 1C, TumorLens** effectively resolves the complex duplication/LoH events affecting HLA class I locus, even when the tumor content is reduced to 90% and 50%. At 90% tumor purity, the pipeline achieved an 86% recall for structural variants, with an additional 10% of the variants being reported as two or more disjoint events. The sensitivity for LoH detection was similarly high at high-to-moderate purities: the pipeline identified 17 and 15 LoH regions at 90% and 70% tumor purity, respectively. However, detection sensitivity declined significantly with increased normal-cell contamination; only one LoH region was detected at 50% purity, and no events were identified below this threshold. We observed a similar trend for cDMRs, with 90% of regions detected at 90% tumor purity, decreasing to 67% at 70% purity and 29% at 50% purity. For HLA typing, TumorLens utilizes a specialized re-mapping strategy (**see Methods**) to assess read-count imbalances distinguishing between genuine LoH and likely erroneous alleles. The pipeline demonstrates remarkable resilience to normal-cell contamination; at 50% tumor purity, TumorLens still successfully identifies 82.7% of the variants. **Figure 1C** shows that a sample with 50% tumor content sample will show a triplication (CN=4 at 100% tumor purity) represented as a duplication (CN=3). Nevertheless, TumorLens is able to report the correct triplication due to an internal calibration when provided with the tumor content *a priori*. **Supplementary Tables 6-8** list all the results for the purity benchmark for samples AOCS21 and AOCS9. Overall, we have demonstrated the ability of the TumorLens pipeline to accurately recapitulate the genomic variants, identify differential methylation patterns and resolve HLA typing at tumor purity as low as 50%.

### Analysis of tumor-normal match tissues samples

Following initial validation, TumorLens was applied to characterize patient-specific mutations and methylation changes in clinical tumor-normal matched-tissue pairs. We sequenced a cohort of 13 lung cancer samples (nine squamous cell carcinoma, two large-cell carcinoma, one adenocarcinoma and one carcinoma of unspecified histology) alongside its corresponding matched normal adjacent tissues. Overall, the ONT sequencing yielded an average coverage of 28.5x per sample with an N50 read length of 16 kb (**Figure 2a**). TumorLens successfully processed all samples in the clinical cohort, identifying multiple types of tumor-specific variants, including somatic mutations and epigenetic alterations (**Supplementary Table 9**). Quality control metrics showed that sample lung_2 likely contained exogenous contamination and was removed from the analysis. We next examined differential methylation patterns across the cohort, analyzing 270,075 genomic regions. While 38.08% of these regions exhibited hypermethylation in at least one sample, we identified a high-confidence “constitutive” signature of 137 regions that were hypermethylated across all 12 tumor samples. Annotation of these regions revealed 114 genes, including transcriptional regulators (*ZSCAN29*, *ZNF561*) and critical mediators of genome stability. Specifically, we identified hypermethylation of *WRAP53*, a p53 regulator linked to poor prognosis in NSCLC, and *TP53BP1*, a central component of the non-homologous end joining (NHEJ) repair pathway. Additionally, epigenetic silencing of *CPT2* suggests a metabolic shift toward altered fatty acid oxidation and immune evasion. Gene Ontology (GO) enrichment analysis corroborated these findings, showing a significant overrepresentation of genes involved in DNA repair, metabolism, and chromosome organization (**Supplementary Table 9**). Together, these results demonstrate that TumorLens can pinpoint a core set of epigenetically inactivated genes that likely drive lung cancer pathogenesis through the disruption of genome maintenance and metabolic homeostasis.

Further analysis of the methylation difference per individual (tumor-normal, see Methods) shows a clear skewing towards higher overall methylation in the normal controls in 11/12 samples (**Figure 2b**), highlighting methylation de-regulation and/or demethylation in the tumor samples, while one sample showed no skewing (lung_8). Given the heterogeneity in tumor subtype and purity among these samples (**Supplementary Table 10**), we performed a high-resolution investigation of each sample independently. We focused on three lung cancer pairs that represent distinct subtypes-represented by individuals lung_3, lung_5 and lung_B-all of which exhibited significant genetic and epigenetic alterations (**Figure 2b**, dotted boxes). More details of the remaining samples are detailed in the **Supplementary text**.

Lung_5 is a tumor sample of unspecified histology that showed 70% tumor content. TumorLens detected 17 large-scale somatic CNVs (**Supplementary Figure 1**) and seven large LoH events (over 10Mb in size). Notably, a 41Mb LoH region on Chr6 was detected that impacts the entire HLA locus. This broad LoH affects a number of genes related to HLA and antigen presentation pathway, including *TAP1, TAP2* and *TAPBP* (Tapasin). The loss of these alleles suggests a reduction in antigen presentation, which has been shown to be associated with higher likelihood of immune evasion, immunotherapy resistance, and shorter overall survival in lung cancer patients^21–23^. The p-arm of Chr9 was also affected by a concurrent deletion and LoH event. This region includes the type I *IFN* gene cluster, its loss suggests a potential for an immune-cold tumor microenvironment. Further, TumorLens reported that 28% of the genome in Lung_5 showed cDMR (see **Methods**). In-depth analysis of the cDMR showed that over 99% of regions have lower methylation levels in the tumor with respect to the matched normal sample (**Supplementary Figure 2**). Here, specific differential methylation was identified within critical immunological loci. These included two HLA class II genes (*HLA-DRA, HLA-DPB1*), four interferon genes (*IFNA4, IFNA16, IFNA17, IFNA1*), seven KIR genes and three T cell-related genes (*TRB*, *TRA* and *CRTAM*). Further, we investigated specific loci showing hypermethylation in the tumor compared to the normal control, as the gene silencing could benefit the tumor progression. **Figure 2c** shows the chromosome-wide methylation difference in Chr9, with the red circle highlighting regions that overlap with the BMP/Retinoic Acid Inducible Neural-Specific Protein 1 gene (*BRINP1*, also identified as *DBC1* (Deleted in Bladder Cancer 1)). This is a tumor suppressor gene that is involved in cell cycle regulation, and has been observed to be frequently silenced by methylation in various cancers such as bladder, ovarian, gastric and non-small cell lung ^24–26^ , suggesting an enhancement in cell proliferation in cancer settings. Additionally, hypermethylation was shown on *PTCSC2* which has been identified as a thyroid cancer susceptibility gene ^27^, likely to interact with transcription factors or modifying chromatin structure.

Finally, the allele-specific methylation (ASM) analysis over 19 *HLA* and 54 APM genes resulted in both *HLA-DQA1* and *HLA-DQB1* showing ASM (**Figure 2d**). By utilizing the normal sample as reference for phasing, we observed LoH in these genes in the tumor (See **Methods**). We were able to discern both alleles in the tumor sample, which was confirmed by the presence of SVs. The second allele in the normal (solid line) showed methylation differences in two regions (highlighted in dotted lines) over an intron of *HLA-DQA1* and an exon of *HLA-DQB1,* the latest clearly showed increase in methylation from a hypomethylated status in the normal control. Furthermore, the first allele (dashed line) is solely present in the normal, with few reads observed in the tumor and HLA typing results confirmed the LoH in the HLA locus (**Supplementary Table 11**). Thus, this targeted phasing approach can aid in the detection of contamination, moreover we do not discard cell heterogeneity or reads from sub-clonal cancer cells.

**Figure 2.**
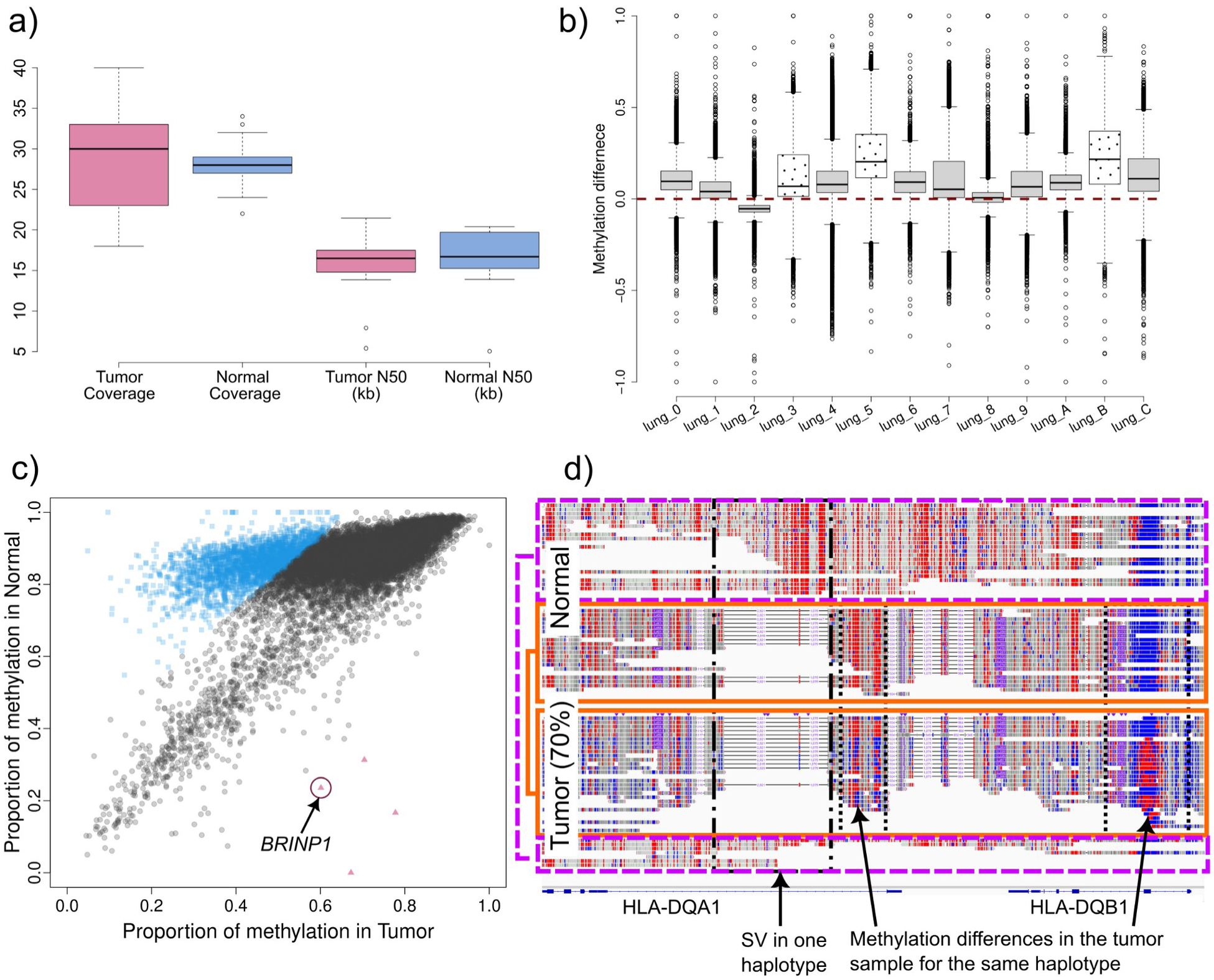
**a)** Coverage and read length statistics for the 13 lung cancer samples with its matched normal adjacent tissues. **b)** Methylation difference between tumor and normal for the 13 lung cancer samples pairs. Genome wide methylation assessment over 10kb non-overlapping windows was performed for each sample and each window was compared to its counterpart in the normal sample. In 11/13 cases, a creak skew towards higher methylation in the normal samples can be seen, with one sample showing no skew and one sample a skew towards the tumor sample. The samples described in the main text are shown with a dotted pattern. **c)** Methylation differences in Chr9 of sample lung_5 (70% tumor purity). Chromosome-wide methylation is shown comparing 10kb non-overlapping windows, with the tumor sample in the x-axis and normal in the y-axis. In blue squares are genomic regions that show higher methylation in the normal sample, while red triangles show genomic regions with higher methylation in the tumor, and gray circles show no significant methylation difference. A clear methylation de-regulation can be seen. Further investigation revealed BRINP1, a tumor-suppressor gene, hypermethylated in the tumor sample. **d)** Allele-specific methylation assessment over HLA-DQA1 and HLA-DQB1. Phasing is done in the normal and transferred to the tumor samples, thus enabling phasing in LoH regions. Allele 1 (dashed line) is solely present in the normal, with few reads from allele 1 observed in the tumor. Allele 2 (solid line) is the one present in the tumor, with the dotted boxes highlighting methylation differences over the same allele in HLA-DQA1 (lower methylation over the intron in the tumor) and HLA-DQB1 (higher methylation over the exon in the tumor).

**Figure 3.**
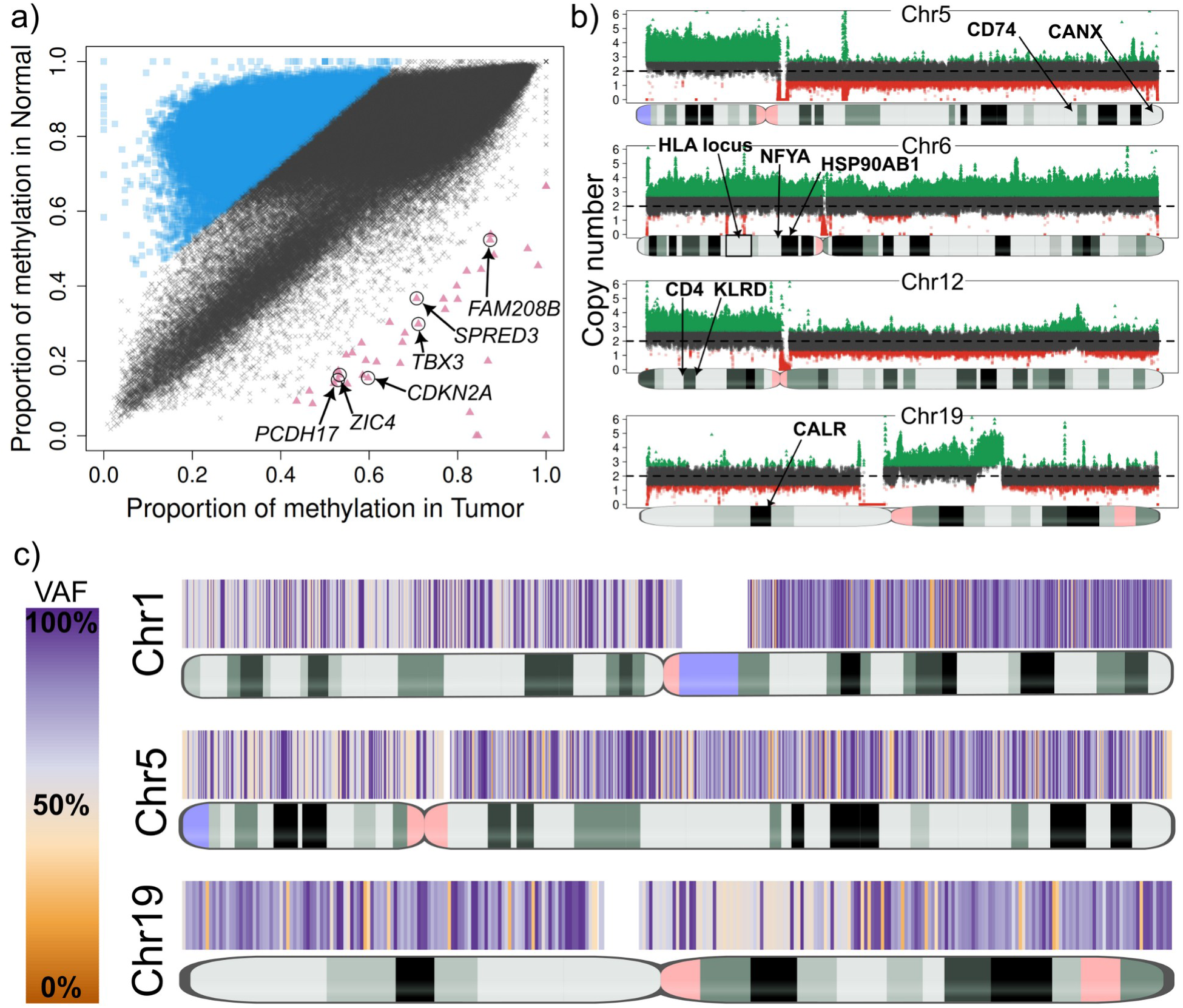
**a)** Genome-wide methylation comparison between tumor (x-axis) and normal (y-axis) in sample lung_B. Blue squares show genomic regions with higher methylation in the normal, which encompass numerous genomic regions with no apparent relation, suggesting widespread methylation de-regulation. The red triangles show genomic regions with higher methylation in the tumor. Curation of these regions revealed six genes with previous reports of silencing in cancer. **b)** Large Copy Number Variants (CNVs) in Chromosomes 5, 6, 12 and 19 affecting the HLA locus and important APM genes in sample lung_B. Coverage plots show deletions and duplications spanning from pieces of chromosomes (DUP in chr19) to whole chromosomes (DUP in chr6). **c)** Loss of Heterozygosity (LoH) heatmap plots of Chromosomes 1, 5 and 19 of sample lung_B. VAF, variant allele fraction.

The lung_B sample is from a patient with large cell carcinoma, showing 65% tumor purity. The sample showed extensive epigenetic dysregulation, with 31% of the genome differentially methylated and overall lower methylation compared to the normal sample (**Figure 3a, Supplementary Figure 4**). Among the global hypomethylation observed in lung_B, a distinct subset of regions exhibited significant hypermethylation. Most notably, TumorLens identified hypermethylation of the *CDKN2A* locus, consistent with silencing of p16(*INK4a*)/p14(*ARF*) tumor suppressors frequently lost in lung cancer^28^. Hypermethylation of additional genes involved in cell adhesion (*PCDH17*) and signaling (*TBX3*) may further enhance tumor progression^29,30^.

Somatic variant analysis of lung_B revealed eight large-scale chromosomal CNVs, including a whole-chromosome 6 duplication (**Figure 3b**) encompassing numerous immune-related genes such as the HLA locus as well as *LTA, TAP1/2, TAPBP, NFYA*, and *HSP90AB1* - all of which are critical for antigen processing and immune regulation. Additional duplications were identified on chromosomes 5p and 12p, the latter involving the KLR gene cluster previously linked to aggressive gastric cancer (**Supplementary Figure 3**). TumorLens further detected eight loss-of-heterozygosity (LoH) regions larger than 20 Mb (**Figure 3c**), spanning *CTSS, CD74, RFX5, CANX,* and *CALR*, which are key mediators of MHC class I/II antigen presentation. The loss of RFX5 impairs MHC class II transcription, while the disruption of *CTSS* and *CD74* hinders peptide loading, collectively restricting T-cell recognition. Loss of heterozygosity in *CANX* and *CALR* may weaken MHC class I peptide presentation, reducing tumor’s immune visibility. Overall, the somatic landscape of lung_B reveals a highly complex immune environment. While MHC class I locus duplications and class II hypomethylation suggest an attempt to maintain antigen display, the defects in antigen-processing machinery offset these effects and may enable immune escape. Moreover, 9p21.3 deletions affecting type-I interferon genes indicate a dampened interferon response, consistent with immune evasion and reduced responsiveness to PD-(L)1 blockade therapy^31^. Together, these findings point to coordinated genetic and epigenetic remodeling that may help the tumor avoid “missing-self” detection while constraining adaptive immune visibility.

The lung_3 sample, a squamous cell carcinoma with 70% tumor purity, displayed extensive epigenetic dysregulation, with 11.1% of the genome classified as differentially methylated regions (cDMRs). Hypermethylation was observed across several transcriptional regulators associated with angiogenesis and proliferation, including *ZNF503*, *GATA2*, and *EPAS1*, all previously implicated in non-small cell lung cancer (NSCLC) (**Figure 4a**)^32–34^. Lung_3 also exhibited signs of localized epigenetic activation within key immune loci. Notably, the interferon gene cluster on chromosome 9p21.3 and *HLA-DQA1* exhibited reduced methylation in the tumor compared to the matched normal sample (T ≈ 0.60 vs. C ≈ 0.93), suggesting activation of interferon signaling and enhanced MHC class II accessibility. Allele-specific analysis revealed asymmetric methylation between haplotypes within the interferon cluster, with up to 43% difference across six *IFNA* genes, consistent with transcriptional allele bias (**Figure 4b**)^35^.

Somatic analysis of lung_3 identified one large deletion and 12 duplications (>10 Mb), including a chromosome 14 amplification encompassing *RFX5* and *CIITA*-the two key regulators of MHC class II transcription (**Figure 4c**). *RFX5* forms an “enhanceosome” complex that protects MHC II promoters from epigenetic silencing, while *CIITA* acts as a master transcriptional coactivator required for MHC II expression. Co-duplication of these genes aligns with a “cancer-cell-intrinsic” MHC II (ciMHC-II) phenotype previously described in NSCLC, which correlates with increased CD4⁺ T-cell infiltration and improved response to PD-1 blockade therapy^36^. Additional duplications on chromosome 19 affected a suite of genes: *CTSS*, *CALR*, *HSPA5*, *PSME1/2*, and *IFI30*, which linked to enhanced antigen processing and aggressive tumor behavior^37–39^. Collectively, the combined hypomethylation at MHC II loci and amplification of *RFX5/CIITA* suggest that this tumor may sustain class II antigen presentation capacity, potentially sensitizing the patient to immune checkpoint therapy.

Across the entire cohort, **TumorLens** demonstrated a high sensitivity for detecting multi-layered genomic instability. This tool resolved widespread epigenetic remodeling with methylation differentials in up to 31% of the genome when comparing tumor and normal (e.g., lung_B, **Supplementary Table 10**). The structural and copy-number landscape across the 13 samples was similarly extensive: four samples showed extensive LoH regions, 10 out of 13 samples exhibited large-scale duplications or deletions exceeding 10Mb) and 10 out of 13 samples harbored large structure variants (>50kb). Notably, 100% of the analyzed samples exhibited at least one major genomic or epigenetic event.

### Analysis of tumor-only samples

Somatic mutations are typically detected via the comparison of a tumor sample to a matched normal control (either adjacent to the tumor tissue or peripheral blood)^3^. However, in many clinical and retrospective settings, matched controls are not always available^40^, presenting a significant barrier to accurate somatic profiling. To enable the ascertainment of tumor-specific mutations, TumorLens deploys automatic filtering and prioritization steps whenever no normal sample is available. This filtering is based on prior computed information across the recently released long read sequencing datasets. To refine SV calling, we utilize STIX^41^, a long-read based annotation tool for SVs, to classify variants as pseudo-somatic if the SV is not present in the STIX database (see **Methods**). For CNVs and LoH events, we utilized strict size filters to prioritize large events (> 1Mb, See **Supplementary Text**). To validate the accuracy of our filtering pipeline, we utilized the ovarian cancer sample AOCS21 to benchmark TumorLens for tumor/normal and tumor-only analysis. The tumor/normal comparison identified 2,294 somatic SVs, while the tumor-only analysis yielded 3,676 pseudo-somatic SVs. When comparing both sets, we found 220 somatic SVs that were absent from the 1000 Genomes Project catalogue. Moreover, 1,958 somatic SVs were actually present within the 1000 Genomes Project catalogue which is based on self-described healthy individuals. For CNV and LoH prioritization, a strict size filter was used (**See Methods**), which resulted in the successful identification of all somatic LoH regions and 63% somatic CNVs (51 out of 81) in ovarian sample AOCS21 using the tumor-only pipeline. For methylation assessment, we implemented a dual-strategy approach to ensure high-resolution detection of cDMRs. 1. per-sample assessment to identify genomic regions as either hypermethylated (>= 90% methylation) or hypomethylated (<= 10% methylation). These extreme states accounted for 30.82**%** of all assessed genomic regions (84,414/273,902). and 2. utilization of a reference panel consisting of 18 blood samples from ten healthy individuals to compare methylation changes^42^. When contrasting matched-normal and panel-of-normals, the tumor-panel comparison resulted in 23.90% (65,473/273,902) of regions as cDMRs, while the tumor-normal comparison resulted in 26.03% (71,299/273,902). Collectively, both results yielded 65,153 shared cDMRs (high recall of 91.38%). Notably, only 320 cDMRs were unique to the tumor-panel comparison, resulting in an exceptional precision of 99.55%.

**Figure 4.**
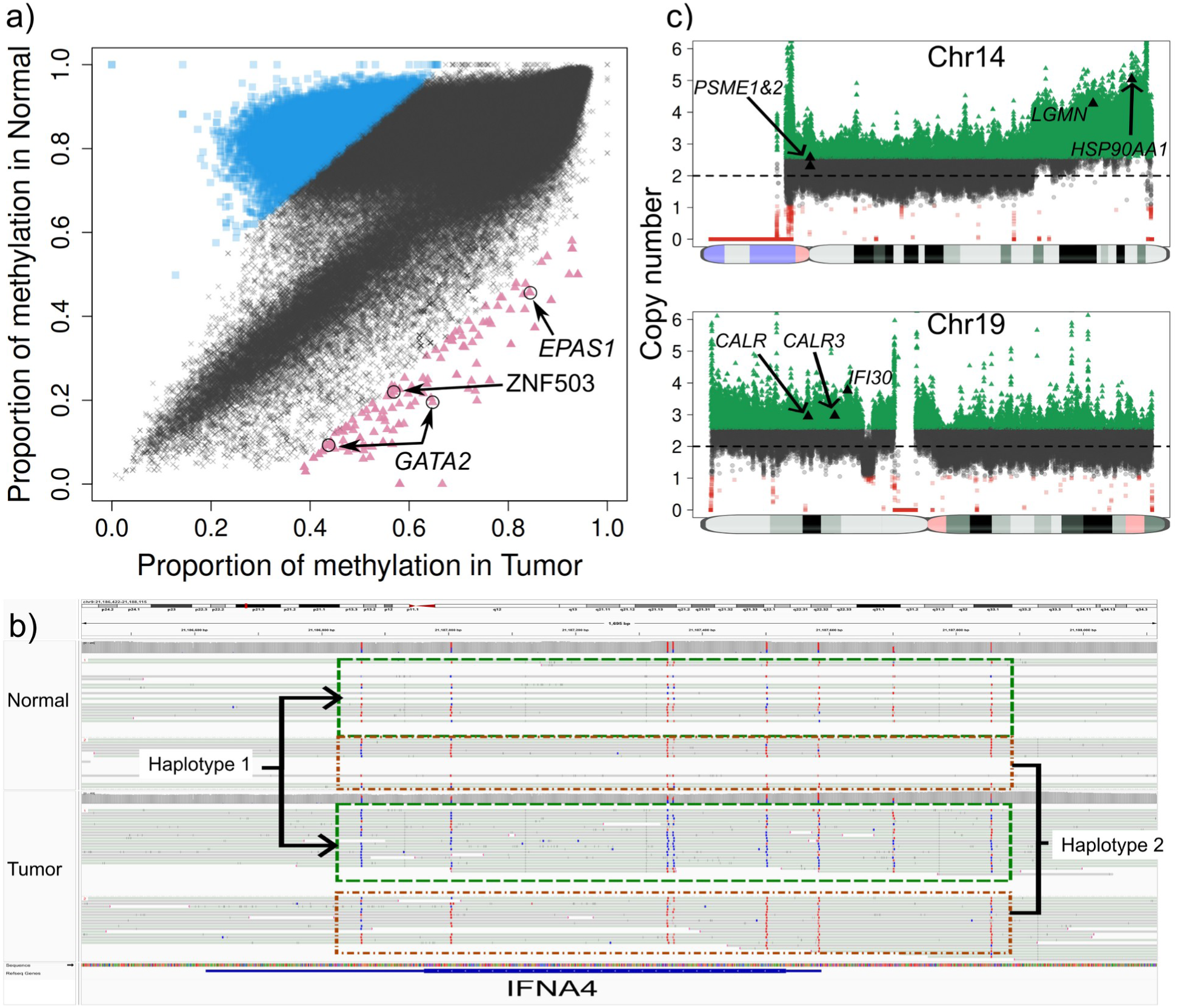
**a)** Genome-wide methylation comparison between tumor (x-axis) and normal (y-axis) in sample lung_3. Blue squares show genomic regions with higher methylation in the normal, which encompass numerous genomic regions with no apparent relation, suggesting widespread methylation de-regulation. Focusing on genomic region with higher methylation in the tumor sample (red triangles) revealed three genes (ZNF503, GATA2 and EPAS1) that have previous reports of being frequently downregulated via methylated or epigenetically repressed in lung tumors. **b)** Allele-specific methylation in the IFNA4 gene in the tumor sample of lung_3. Haplotype one is shown as differentially methylated when compared to the same haplotype of the normal sample. Allele-specific methylation in cancer can be disruptive in transcription factors. **c)** Large-scale CNVs (duplications) in Chromosomes 14 and 19 which affect several genes including seven genes involved in the antigen presentation pathway.

To further validate the pipeline’s performance in a real-world clinical scenario, we analyzed a cohort of stomach cancer samples where matched normal tissue was unavailable. We sequenced five stomach tumor-only samples of unspecified histology and tumor purity (coverage: 24-29x) and analyzed them with TumorLens. Across the cohort, we detected an average of 23,332 total SVs per sample , from which roughly 450 unique pseudo-somatic SVs per patient were identified based on the population annotation by STIX (**Supplementary Table 12**). Furthermore, our prioritization analysis of CNV and LoH produced an average of 5 high-confidence CNV regions and 9 LoH regions per sample (**Supplementary Table 12**).

Further, methylation analysis was performed using the per-sample strategy (highly and lowly methylated regions). All samples exhibited at least 9% of genomic regions as highly methylated (**Supplementary Table 12**). Interestingly, while the broader genome underwent extensive remodeling, the 74 HLA and APM genes remained strikingly stable-none of these critical immune-recognition genes were classified as highly or lowly methylated. A deeper investigation into all protein coding genes showed that 7,894 genes were hypermethylated, of which 7,846 genes (99.4%) were universally hypermethylated in all five samples. To understand the biological consequences of this universal silencing, we performed a Gene Ontology analysis on the shared hypermethylated gene set. The analysis revealed nine overrepresented cell-processes, all of which were related to the immune response (**Supplementary Table 13**).

By contrasting our stomach samples against the reference panel of 18 healthy blood samples, we identified high-confidence hypermethylation events at several critical loci: genes *PARM1, CDKN2A* and *CAT* showed highly methylation in the tumor sample stomach_1 (**Figure 5a**). These genes have been reported to be often silenced (by methylation) or repressed in cancer^43–45^. Next, in the tumor sample stomach_3, *SLX4IP* was identified as highly methylated. This gene regulates telomere dynamics and its inactivation can affect metastasis and patient survival^46^. Further, in sample stomach_2, we detected 321 genes with higher methylation in the tumor. A Gene Ontology analysis showed that many genes relate to processes of both blood cell differentiation and the digestive track. Moreover, manual curation of the hypermethylated loci found six genes: *MECOM, NF1, FRZB, EOMES, ALDH1A2* and *SOX17* in which loss or reduced expression (often by promoter hypermethylation) has been observed in various cancers^47–49^. Finally, we focused on “constitutive” hypermethylation events - regions that were consistently highly methylated across all five stomach cancer samples but remained hypomethylated in the panel of normals. TumorLens detected six specific regions affecting four genes that exhibited this tumor-specific hypermethylation pattern. Among these, *GPR21* is a particularly significant finding, our results align with emerging evidence that *GPR21* expression is significantly reduced in various tumor tissues including cervical, breast, skin, prostate, as well as astrocytoma when compared with healthy fibroblasts ^50^.

To conclude the gastric cancer cohort analysis, we examined the structural and copy number landscape. Despite the lack of matched controls, **TumorLens** identified high-confidence structural events that mirror the immune-evasion strategies observed in our lung cancer cohort.

In sample Stomach_2, we detected a likely somatic 928Kb heterozygous deletion that affected a cluster of five interferon genes (*IFNA6, IFNA13, IFNA2, IFNA8, and IFNA1,* **Figure 5b**). This deletion is a critical finding, as the loss of Type I interferon genes can disrupt the normal immune response, and has been suggested that it could lead to resistance to immunotherapy based on *in-vitro* analysis ^51^. Simultaneously, CNV analysis detected duplications of several genes essential for antigen presentation: *CALR,* a chaperone involved in MHC class I assembly; *IFI30, an enzyme* crucial for MHC class II antigen processing and *RFXANK, a core subunit of the RFX complex required for MHC class II transcription.* Then, in sample stomach_3, TumorLens detected a DUP/LoH complex event on Chr8 affecting the *CTSB* gene (**Figure 5c**, with copy number = 7). The upregulation of CTSB has been reported in certain cancers and premalignant lesions^52,53^. Finally, HLA typing across the gastric cohort provided further evidence of selective pressure on the immune-recognition machinery. While true LoH is difficult to definitively confirm without a normal sample, the high frequency of homozygous HLA typing strongly hints at somatic loss events: a single class I gene *HLA-B* exhibited homozygosity in sample stomach_4; class II genes showed homozygous typing in *HLA-DPA1* and *HLA-DPB1* for samples stomach_1, _2 and _3; *HLA-DPA1* in sample stomach_4 and *HLA-DQA1* and *HLA-DQB1* for sample stomach_5 (**Supplementary Table 14**).

Overall, we have demonstrated that our variant prioritization methods, when coupled with advanced gene annotation, effectively resolve the genetic and epigenetic landscapes of tumor-only samples.

By utilizing the unique capabilities of Oxford Nanopore long-read sequencing, TumorLens consolidates the detection of structural variants, copy number alterations, and DNA methylation into a single, unified assay. This integration eliminates the need for multiple disparate technologies, providing researchers with a high-resolution, phased view of the cancer genome. Whether identifying rare somatic structural variants or universal epigenetic silencing signatures, **TumorLens** bridges the gap between raw sequencing data and deep biological insight, even in the most challenging clinical contexts where matched controls are unavailable.

**Figure 5.**
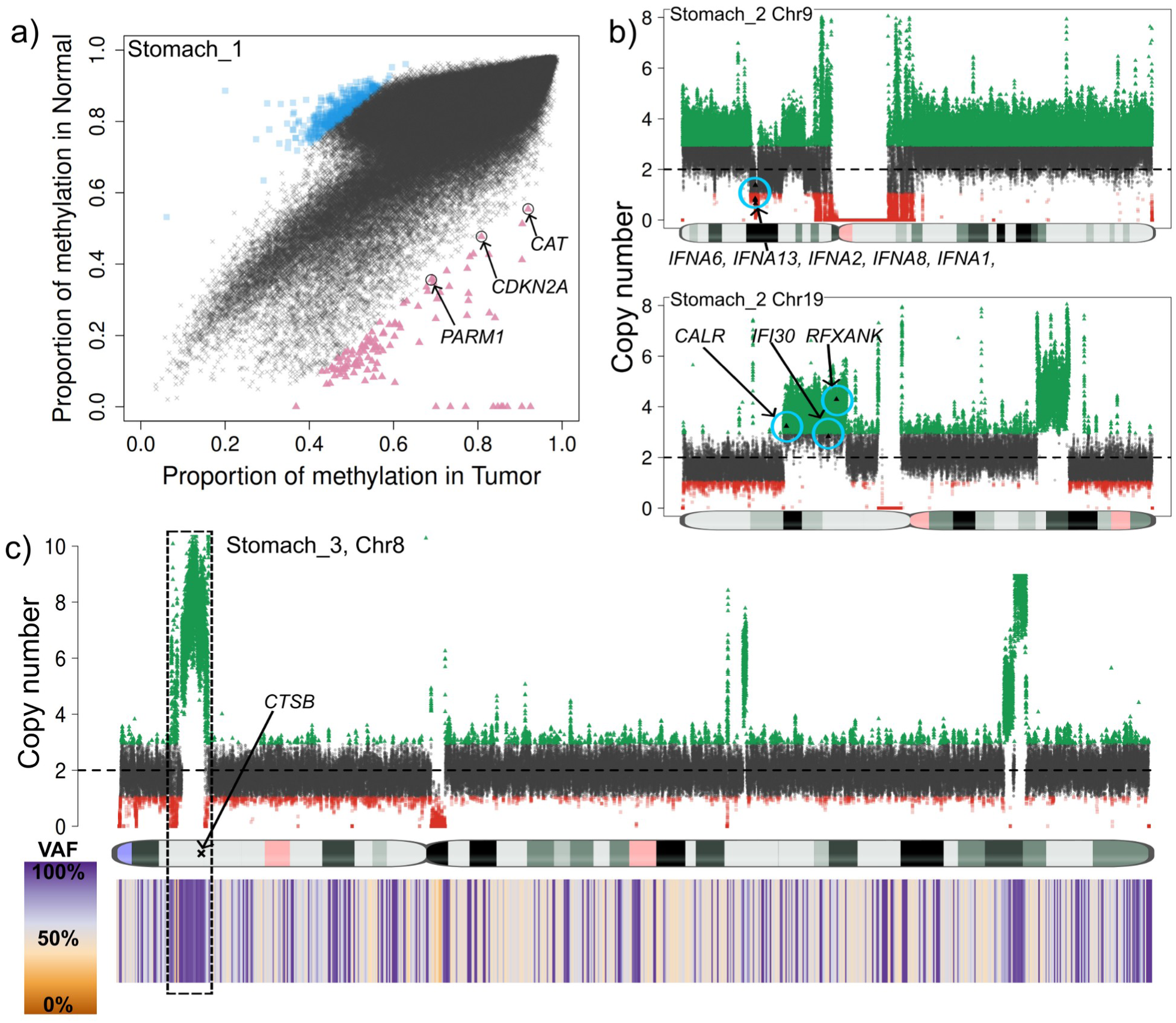
**a)** Genome-wide methylation comparison between tumor (x-axis) and the panel of normals (y-axis) in sample stomach_1. Blue squares show genomic regions with higher methylation in the panel of normals while the red triangles show regions with higher methylation in the tumor sample. Gene annotation of the high methylation in the tumor regions reveals genes PARM1, CDKN2A and CAT which have been reported to be often silenced (by methylation) or repressed in cancer. **b)** Deletion (top) and duplication (bottom) detected in the tumor sample stomach_2. The deletion was detected as an SV and curated with STIX as not being present in the 1000 genomes, and thus classified as pseudo-somatic. This deletion affected five genes from the IFN gene family. 7.85Mb duplication detected by CNV in the p-arm of Chr19. This duplication affects CALR, which is involved in MHC class I antigen presentation, IFI30 and RFXANK which play a role in MHC class II expression and function. **c)** Complex 4Mb long DUP/LoH (CN=7) detected in the p-arm of Chr8 in sample stomach_3. This DUP/LoH affects CTSB which has been reported as upregulated in certain cancers and premalignant lesions.

## Discussion

Here, we present TumorLens, an integrated novel computational pipeline for long-read sequencing that allows the concurrent, genome-wide analysis of genetic (SNV, SV, large CNV, LoH) and epigenetic (5mC methylation) variation. By incorporating a personalized reference approach, TumorLens improves the ascertainment of HLA types and LoH status while providing easy-to-interpret summary reports and figures that allow non-expert users to interpret tumor-specific mutations, even in tissues with variable tumor purity (down to 50%). We validated the accuracy of TumorLens across three benchmark cell lines and demonstrated its utility in a total of 15 additional cell-lines and clinical tumor/normal tissue pairs. Furthermore, we showed that TumorLens remains robust in “tumor-only” scenarios-analyzing five gastric cancer samples with unknown purity estimations-thereby enabling the accurate delineation of somatic landscape in both prospective and legacy clinical cohorts.

TumorLens leverages the unique capability of long-reads to capture methylation and DNA sequencing data on a per-molecule basis, without the need of bisulfite conversion or other biochemical alterations on the DNA itself. Although only tested for ONT sequencing, the pipeline remains technology-agnostic. The length of the reads significantly improved the classification of variant types (e.g Structural Variants) and mappability in complex, repetitive regions such as the HLA locus. While similar integrative methods exist for short-read sequencing, the consolidation of these multi-omic layers is novel in the long-read space and the presented methods could be extended beyond the tumor-normal comparison to other tissue comparisons (e.g. in the SMaHT network^42^). Moreover, we demonstrate that by leveraging population-scale control data-such as the 1000 Genomes Project ^41^ and the SMaHT network-TumorLens can accurately prioritize tumor-specific somatic variants and methylation abnormalities in tumor-only analysis. This predictive power will continue to grow as more long-read data becomes available from multiple large scale studies, which can be utilized as important reference points^42,54,55^.

Cancer studies that analyze only one variant type, such as CNVs or SNVs, overlook the reality that genetic and epigenetic alterations coexist and interact on the same molecule to drive tumor evaluation. Our multi-layered analysis highlights this interplay. For example, the duplications of *RFX5* and *CIITA* coinciding with *HLA-DQA1* hypomethylation suggest a coordinated program to sustain MHC class II expression, while *CDKN2A* hypermethylation and interferon locus loss illustrate concurrent genetic and epigenetic silencing of immune pathways. Such patterns emphasize that tumor evolution and immune escape cannot be fully understood without integrating structural, sequence, and epigenetic variation at single-molecule resolution.

Despite these advantages, our approach has several limitations. First, HLA typing accuracy, while high, currently remains below what is achieved with short-read NGS-based methods, which can exceed 99% concordance at the two-field level^56^. Second, tumor purity below 50% continues to pose challenges for variant detection and methylation calling, as low tumor content dilutes somatic signals and complicates haplotype phasing. Future work that focuses on enrichment of relevant genomic loci using adaptive sampling ^57^ may help overcome these constraints and further enhance the accuracy of TumorLens in highly heterogeneous samples.

Our understanding of tumor development and progression continues to expand with advances in sequencing technology. Until recently, parallel assessment of genetic and epigenetic alterations required separate library preparations, often hindered by tumor heterogeneity and limited yield. This challenge has driven the rise of single-cell and spatial transcriptomic approaches. Tools such as TumorLens can enable detection of somatic and mosaic mutations but remain limited by coverage, reducing sensitivity for variants below ∼5% variant allele fraction and for nonsense mutations. In contrast, our approach combines comprehensive variant detection with clinically relevant turnaround times. As demonstrated previously ^58^, we can generate and analyze 60× whole-genome data within 6-8 hours of sample collection, and recent studies show that long-read–based analyses can even be performed intraoperatively. Such rapid, integrated genomic and epigenomic profiling holds significant promise for more accurate diagnoses and real-time, personalized treatment decisions.

In conclusion, TumorLens harmonizes the analysis of all major genomic and epigenetic modifications into a single, cohesive framework. By resolving the hidden complexities of the cancer genome, TumorLens provides a scalable pathway toward improving both the diagnostic depth and clinical turnaround times for precision oncology.

## Methods

### Sample selection

#### Cell lines

Patient-derived cell lines H2009 (CRL-5911) and BL2009 (CRL-5961) were established from tumor and matched normal tissues of a lung cancer patient, obtained from ATCC.

Patient-derived cell lines AOCS9 ^59^, AOCS21 ^60^ and AOCS19 were established from ascites collected from ovarian cancer patients recruited to the Australian Ovarian Cancer Study (AOCS), an Australian multi-center study with ethics approval from the Peter MacCallum Cancer Centre ^61^. Approximately 2 mL of ascites fluid collected at recurrence was centrifuged at 1500 rpm for 5 min to create a cell pellet. Supernatant was removed and the cell pellet was resuspended in 10 mL of complete RPMI media (RPMI 1640, 10% FBS, 50 µ/mL penicillin and 50 mg/mL streptomycin) and transferred into a standard humidified incubator (37 °C, 5% CO_2_). Media were replaced after 48 h, and then once every 2-4 days, until an adherent cell line was established. Cells were passaged ten times from the time of collection and stocks cryopreserved. All cell lines were authenticated against the patient germline DNA using STR profiling (GenePrint 10 System, Promega).

#### Clinical samples

Five stomach tumor tissues were obtained from ProteoGenex. Thirteen matched pairs of lung tumor/NAT (normal tissue adjacent to the tumor) were obtained from University of Michigan, ProteoGenex or SeraCare LifeSciences. Each provider received IRB approval of research, appropriate informed consent of all subjects contributing biological materials, and all other authorizations, consents, or permissions as necessary for the transfer and use of the biological materials for research at Genentech.

#### HLA LoH statuses for selected samples

HLA LoH statuses for the cell lines were identified using a combination of short read sequencing technologies and computational HLA genotyping.

Whole genome sequencing (WGS) was performed for NCI-H2009 (H2009) and NCI-BL2009 (BL2009). WGS libraries were generated from 100ng of genomic DNA using the Illumina DNA Prep kit (Illumina). Libraries were pooled and sequenced on NovaSeq 6000 (Illumina) to generate 1-billion paired-end 75-base-pair reads for each sample.

Whole genome sequencing (WGS) was performed for AOCS9, AOCS21 and AOCS19 germline and cell line pairs. WGS libraries were generated from 1 μg of genomic DNA using TruSeq DNA PCR-free sample preparation protocol (Illumina, San Diego). Sequencing on a HiSeq X Ten (Illumina at GenomeOne, Kinghorn Comprehensive Cancer Centre, Sydney Australia) was performed to a minimum average of 30-fold base coverage for germline and cell line DNA.

Using HLA-HD ^62^ v1.7, computational HLA genotyping for HLA I genes HLA-A, -B and -C was performed on the WGS of the selected samples. Subsequently, we compare the HLA-A, -B and -C alleles between each tumor/normal pair to determine whether there is HLA LoH. Specifically, for each HLA I gene, if we observe heterozygosity in the normal sample, and homozygosity in the tumor, i.e. one of the alleles in the same gene from the normal has been lost in the tumor, the gene is considered to have HLA LoH. We do not consider the HLA I gene to have LoH if homozygosity is observed in both the normal and tumor samples. We did not observe instances where, for the same HLA I gene, allele homozygosity is observed in the normal and heterozygosity in the corresponding tumor sample.

### Long read sequencing

#### DNA extraction

All selected cell lines were maintained at 37°C in 5% CO2. The H2009 and BL2009 cell lines were grown in RPMI1640 media supplemented with 10% FEB, and 2mM L-glutamine. The ovarian cell lines AOCS9, AOCS21 and 34984 were grown in RPMI1640 media with 10% FBS. DNA was extracted using the Nanobind CBB kit (PacBio catalog number 102-301-900) and stored at 4°C until use.

All clinical samples used in this study were flash-frozen tissues. Genomic DNA was extracted using either the Nanobind tissue kit (PacBio, catalog number 102-302-100) or Monarch HMW DNA extraction kit for tissue (New England Biolabs, catalog number T3060) and stored at 4°C until use.

Genomic DNA was quantified in triplicate using the Qubit dsDNA BR Assay Kit (Thermo Fisher Scientific, catalog number Q32853). DNA purity was assessed using Spectrophotometer (DeNovix DS-11 FX). DNA quality was assessed using Genomic DNA ScreenTape (Agilent Technologies, catalog number 5067-5365) on TapeStation4200 (Agilent Technologies). To increase nanopores occupancy and flow cell output, 2-2.5 µg of genomic DNA was fragmented prior to library preparation by centrifuging in Covaris g-tube (Covaris, catalog no. 520079) at 1300 x g for 1 min followed by inverting the g-tube for 1 minute. The size and quality of sheared DNA were checked using Genomic DNA ScreenTape on TapeStation4200. The final genomic DNA fragments centered at 40-58 kb.

#### Library prep and sequencing

Libraries were prepared from 1.5-2 µg of genomic DNA using the Ligation Sequencing Kit V14 (Oxford Nanopore Technologies, SQK-LSK114). 220ng of library from each sample was sequenced on a R10.4.1 PromethION flow cell (FLO-PRO114M) for 72 hours. For some libraries, washing and reloading flow cells 1-2 times using the Flow Cell Wash Kit (EXP-WSH004) and Sequencing Auxiliary Vials V14 (EXP-AUX003) during their sequencing runs were performed to maximize data output. Raw sequencing data including methylation status was basecalled using Guppy 6.2.11, 6.3.9 and 7.0.9.

### TumorLens

The TumorLens pipeline is implemented in python (tested in 3.10, 3.11 and 3.12) and it is composed of two main steps: Sample processing and somatic analysis.

#### Sample processing

The sample processing step features a highly parallelized pipeline that takes sequence data generated by ONT PromethION machines in either FASTQ or BAM format. Each input file contained both the sequencing and the methylation data, the latter was presented via the MM/ML tags in BAM files and encoded within the sequence name for the FASTQ files.

##### Alignment of long reads

We used Minimap2 ^63^ (version 2.24-r1122) to perform the long reads alignment to human genome version GRCh38.p13 with removed alternative contigs. Minimap2 was run utilizing the Oxford Nanopore preset (-x map-ont) and generating output in SAM format (-a). Subsequently, we converted the alignment to BAM format, sorted, and indexed using SAMtools ^64^ (version 1.19). Following read alignment, TumorLens initiated parallel tasks to analyze genomic variations, compute coverage and profile methylation. Depending on the computational infrastructure configurations, these processes can be parallel or sequential.

##### Coverage computation

Genome-wide coverage was computed using mosdepth ^65^ (version 0.3.8) using a window size of 1 kilobase (kb, --by option), skipping the output per-base depth (--no-per-base) and using only reads with a mapping quality greater-equal to 10 (--mapq 10).

##### Identifying genomic variations (SV, CNV, SNV)

The pipeline performed genome-wide calling of both large and small variants. Small variants, specifically Single Nucleotide Variants (SNVs), were called using Clair3 ^66^ (version 1.0.10) for the ONT platform (--platform=“ont”) with the appropriate model (--model_path). To improve run-time, the genome was split into 20 megabases (Mb) chunks and SNVs were called on each chunk. Once all chunks were processed, they were combined using BCFtools concat (version 1.19,^64^), allowing overlaps (--allow-overlaps) and removing duplicate calls (--remove-duplicates). Finally, BCFtools was used to filter all the SNVs (--apply-filters PASS), only keeping SNVs with an allele fraction greater than 10% (--type snps --include “AF > 0.1”). For tumor-normal pairs, we performed a two-step phasing workflow. First, the SNVs called from the normal sample were phased using WhatsHap^67^ v2.3 with default parameters. Next, the tumor sample was phased using the normal phasing as a reference backbone. This strategy ensured the tumor sample can be phased even in LoH regions. Structural Variants (SVs) were called using Sniffles ^9^ (version 2.6.3) with default parameters, including the reference genome (--reference) and including the SNF output (--snf). Additionally, Sniffles 2 was also run in mosaic mode (--mosaic) to detect low frequency variants (5-22%). Finally, CNVs and LoH were assessed using Spectre ^68^(version 0.2.2-cancer-250529, available at https://doi.org/10.5281/zenodo.18806405). This modified version used tumor/normal pairs to assess somatic CNVs as follows: spectre Cancer --coverage tumor.bed.gz normal.bed.gz --snv tumor_snv.vcf.gz normal_snv.vcf.gz --output-dir outdir_cnv/ --reference grch38.fa --metadata grch38.dmr --blacklist grch38_blacklist.bed --tumor-content {tumor_content} --ploidy 2, where {tumor_content} was the estimated tumor content between 0 and 1. Please see the **Supplement text** for additional details. Finally, for the tumor only samples, Spectre was run in default mode including SNVs as follows: spectre CNVCaller --coverage tumor.bed.gz --snv tumor.vcf.gz --output-dir outdir_cnv --reference grch38.fa --metadata grch38.dmr --blacklist grch38_blacklist.bed

##### Methylation profiling

We extracted genome-wide methylation signals using modkit pileup (version 0.5, https://github.com/nanoporetech/modkit) and then summarized the results into 10kb windows using modkit stats, resulting in 270,075 assessed genomic regions across the genome. Additionally, we used 13 lung normal samples to create a distribution of expected methylation change by performing all pair comparisons among the normal samples. Combined across all pair comparisons, we analyzed 21,416,679 regions in total and asked at which methylation difference threshold do we expect to observe ∼50 candidate regions for the same tissue sample of different individuals, which resulted in a methylation difference threshold of ±33 percentage points. For tumor-normal samples, we assessed genome-wide cancer differentially methylated regions (cDMRs) between tumor and paired normal samples using modkit dmr, assessing only 5mC modifications at CpG motifs (--cpg), to then, based on phasing information we splitted the bam files overlapping the 74 genes listed in (**Supplementary table 1**) with 2kb padding and performed allele-specific differential methylation analysis with methylation modkit dmr in a 4-by-4 way (all-vs-all). For the case of tumor-only samples, we utilized the summarization of the methylation profile of 18 blood samples corresponding to 10 donors from the SMaHT project. We took the proportion of methylated CpGs over 10kb non-overlapping windows and used the median value across all 18 samples as a normal control to assess cDMRs.

##### HLA-specific processing

In addition to the genome-wide analysis, we performed a targeted analysis on the HLA locus, defined as the chromosomal coordinates between 26 Mb and 36 Mb on human Chr6 (chr6:26,000,000-36,000,000). First, we extracted all reads overlapping the aforementioned region and performed HLA typing with specHLA (version 1.0.3, ^69^) using the long reads option. Next, we reduced the typing results to two fields, given that long-read typing is not as accurate as short-read-based typing. Using the assigned types in the normal, we re-mapped the reads of both the normal and tumor to the best hit representative of the given HLA-type (i.e HLA-A*01:01 and HLA-A*24:02) using minimap2 (minimap2 -ax map-ont -t 4 {hla_gene_fasta} {reads}) and counted the number of reads overlapping each HLA type. The ratio difference between the normal Allele1:Allele2 to the tumor Allele1:Allele2 is used to assess the likelihood of LoH.

#### Somatic analysis

##### Paired samples

For each paired-sample, we ran TumorLens and collected genome-wide SVs, CNVs, LoH and methylation data of both the tumor and normal samples. For the SV data, we used Sniffles2 merge to assess cancer somatic SVs, as those present only in the tumor sample (sniffles --input normal_sv.snf tumor_sv.snf --vcf merge_sv.vcf.gz --reference grch38.fa). For CNVs, spectre cancer has a built-in comparison somatic CNV and LoH analysis which is outputted as VCF. The genome-wide differential methylation analysis was done over 10kb windows non-overlapping windows using modkit dmr -a {control} -b {tumor}.

Next, we assessed 74 immune-related genes (**Supplementary Table 1**), which include the HLA gene locus. First, phased SNVs were used to haplotag the bam file over these 74 genes. We also assessed if any of the somatic SV and CNV overlap with any of the genes with a padding of 2kb downstream and upstream to include the promoter regions. The reads overlapping these genes were extracted and haplotagged based on the phased SNVs. Subsequently, each haplotype was split for independent methylation profiling to assess allele-specific methylation in a 4-by-4 way. Additionally, we collected the typing results of 8 HLA genes (*HLA-A, HLA-B, HLA-C, HLA-DPA1, HLA-DPB1, HLA-DQA1, HLA-DQB1, and HLA-DRB1*) and compared the typing results of tumor and normal, taking as ground truth the normal sample results and used the HLA re-mapping results to improve LoH analysis over the HLA genes.

##### Tumor only samples

For each tumor-only sample, we ran TumorLens and collected genome-wide SVs, CNVs, LoH and methylation profiling over 10kb windows and HLA typing. Variant prioritization was performed using specific filters for each data type. For SVs, we used STIX with 1,108 genomes from the 1000 Genomes Project (1KGP) and only assessed those variants absent from the 1KGP index. We only analyzed calls exceeding 1Mb size for CNVs and LoH regions.

### Benchmark

#### SV benchmark

The HG002-Q100 v1.1 GIAB benchmark was used to evaluate Sniffles performance in somatic calling. First, we assessed all SVs in HG002 by extracting the HG002 columns from the merged VCF and comparing them to the benchmark using Truvari v5 as follows: truvari bench –base {base_VCF} --comp {comp_VCF} --output hg002_bench --passonly --includebed giab_grch38.bed --refdist 1000 --reference giab_grch38.fasta. For the case of somatic SVs, the support vector (SUPP_VEC) tag in the info field was used to extract SVs uniquely present in HG002, which was also benchmarked to the GIAB benchmark using Truvari v5 with the same parameters.

#### CNV benchmark

The HG008 GIAB CNV benchmark (v0.4, dated 2025.07.17) was used to assess Spectre performance in somatic CNV calling. We assessed all CNVs larger than 100kb as Spectre Cancer only reports CNV events of 100kb and above. Spectre Cancer reports both somatic and non-somatic variants, so for the comparison we only included somatic variants using bcftools as follows: ‘bcftools view –include “SOMATIC = 1” {vcf} | bgzip -c > {vcf_somatic}’. The comparison was performed with bedtools using two bed files: GIAB CNV bench as base and the output of Spectre as comp using 70% minimum overlap of the benchmark CNV and showing the comp match as follows: ‘bedtools intersect -a {giab_cnv_pass_bed} -b {spectre_cnv_somatic} -f 0.7 - wao’. Missing IDs from the matching base-comp CNVs were used to compute FP and manual inspection of the FN showed split and overlapping events.

#### LoH benchmark

The H2009 lung cancer cell line (NCI-H2009) was used to assess LoH detection accuracy. The NCI-Navy Medical Oncology Branch cell line database reported LoH in several lung cancer samples, including H2009, based on known microsatellites.

## Data Availability

All data produced in the present study are available upon reasonable request to the authors

https://epi2me.nanoporetech.com/giab-2023.05/

https://ftp-trace.ncbi.nlm.nih.gov/ReferenceSamples/giab/data_somatic/HG008/Liss_lab/UCSC_ONT_20231003/

## Acknowledgements

LFP is funded by the NIH grant UM1DA058229. FJS and YF are funded by the NIH grant UG3NS132105.

The Australian Ovarian Cancer Study (AOCS) was supported by the U.S. Army Medical Research and Materiel Command under DAMD17-01-1-0729, The Cancer Council Victoria, Queensland Cancer Fund, The Cancer Council New South Wales, The Cancer Council South Australia, The Cancer Foundation of Western Australia, The Cancer Council Tasmania and the National Health and Medical Research Council of Australia (NHMRC, ID400413 and ID400281). We acknowledge the vital role of the Australian Ovarian Cancer Study and for this study. AOCS acknowledges additional support from Ovarian Cancer Australia and the Peter MacCallum Cancer Centre Foundation, and acknowledges the cooperation and contribution of the participating institutions in Australia and study nurses, research assistants and all clinical and scientific collaborators including Nadia Traficante, Sian Fereday, Linh Nguyen, Joy Hendley, Leanne Bowes and Kathryn Alsop. The complete AOCS Study Group can be found at [www.aocstudy.org].

