## Supplementary text for "Comprehensive detection of genetic and epigenetic alterations in cancer using long reads with TumorLens"

Supplement

### Supplementary notes

#### Spectre cancer

Spectre’s cancer mode is a fork of the Spectre CNV caller that integrates tumor content and on-the-fly merging to assess somatic CNV in tumor-normal pairs. Unlike vanilla Spectre, it requires coverage and SNV calls for both tumor and normal samples. First, it performs the candidate search of the normal sample and utilizes SNVs to assess the coverage for the neutral ploidy (default = 2). Here, Spectre uses “perfect-heterozygous” sites which are SNV which VAF is for diploid organisms, 0.5 +- 0.12, from which average coverage is estimated alongside thresholds for the deletion and duplication search and LoH assessment. Next, it performs the candidate search of the tumor sample, again using the SNVs to assess the coverage for the neutral ploidy. Here, the definition of the thresholds for deletion and duplication search is modified by the tumor content, such that we assume 2x is added as a known proportion to any given genomic region. So if a region is 3x with a 80% tumor content, we expect the average coverage to be 2.8x. Additionally, for LoH assessment the threshold of homozygosity is modified based on the tumor purity, by assuming the normal sample will only add heterozygous sites at with VAF=50% as follows: let us assume the threshold for homozygosity is a VAF = 75% for a 100% tumor content sample, then a 80% tumor content sample with expect the homozygosity is a VAF = 70%. In practice, both the homozygosity VAF and the heterozygous VAF are computed from SNVs of the normal sample. Then, Spectre will output CNV for the normal sample. Finally for the cancer sample it will take the in-memory candidates for the normal sample and compared them to the tumor to assess somatic CNV calls on-the-fly, and will add a “SOMATIC” flag to the INFO field of the VCF output of the tumor sample whenever a CNV or LoH call is only detected in the tumor sample.

#### HG002 benchmark

The HG002/HG003 samples were used to assess both HLA typing and somatic structural variants (i.e. 50bp or larger insertions and deletions), with the SVs shared between HG003 and HG002 considered germline and the HG002 unique SVs as pseudo-somatic. Our pipeline detected the HG002 Genome in a Bottle (GIAB) Q100 structural variants (SVs) with 95.2% precision and 76.9% recall (F1 score = 84.5%). Among these, 6,340 SVs were classified as somatic, with a precision of 89.8%. Next, we assessed HLA typing by comparing the HG002 HLA types for six HLA genes between TumorLens assessment and GIAB benchmark ^1^. **Supplementary Table 3** shows the typing results compared to the benchmark at two-field level. Overall, we correctly identify 12/12 alleles at one-field and 10/12 alleles at two-field resolution. Finally, TumorLens also reported two LoH regions in Chromosomes 1 and 10 (3.5 and 2.9 Mb respectively)

#### AOCS

Sample AOCS9 presents 30 somatic LoH regions, including whole Chr6 LoH (detected both by CNV and typing) and three being larger than 100Mb in Chr 4,10,11 (**Supplementary Table 6**). Further, TumorLens detected a high number of copy number alterations (CNA) in almost all chromosomes. For the case of large SVs (over 10kb) TumorLens detected 52 DEL, 65 DUP, 17 INS, and 44 INV. Further, we detected 55,578 Somatic SNVs.

The sample AOCS19 presents 12 somatic LoH regions including the whole interferon gene cluster. For Structural Variants (SVs) we detected 59 very large somatic SVs, which include a 170Mb deletion in chromosome 1 and a 74Mb deletion in chromosome 14 (average size of SV > 10kb is 13.8Mb, **Supplementary Table 6**), partial deletion of *CTSS*, *RFX5* genes and complete deletion of *PSME1*, *PSME2*. Alongside these, we detected seven large somatic CNVs and 53,118 somatic SNVs.

#### Assessing cancer samples with low tumor content

Since tumor samples rarely consist solely of cancer cells, it is imperative to consider tumor purity-the proportion of cancer cells in a sample-to ensure accurate analysis.

The presence of non-cancer cells reduces the tumor content and makes it harder to identify variants and methylation changes. TumorLens is designed to address these challenges. Briefly, TumorLens takes into account the non-cancerous control sample (hereafter referred as normal) contribution to automatically adjust the VAF utilized for LoH and SV detection, corrects the duplication and deletion definition for CNV and accounts for the reads of the normal sample by phasing before differential methylation analysis (see **Methods**). To demonstrate this across variant and methylation calls, we mixed the ovarian samples AOCS21 and AOCS9 to simulate tumor content levels ranging from high (90%) to low (10%). **Supplementary Tables 6** shows the changes in detection of methylation differences, somatic SV, somatic CNV and LoH regions (**Figure 1C**), while **Supplementary Tables 7** and **8** describe typing results for the same titration experiments.

In all the analysis, the decrease in tumor content closely correlated with decreased detection of methylation differences and somatic variants. First, we examined genome-wide differentially methylated windows. At tumor contents of 70% and above, we were able to detect at least 50% of the differentially methylated regions identified in the pure tumor samples. At 30% and lower tumor contents, we only detected 2% of the regions that showed methylation differences in the pure tumor sample. Thus, detecting hypermethylation increases in difficulty with lower tumor concentrations. To mitigate this effect, TumorLens utilizes phasing information to perform haplotype differential methylation, in an attempt to reduce the signal dilution. For example, comparing the methylation proportion over *HLA-A* in sample AOCS9 shows the methylation for the normal sample being 7.41% for A*23:01 and 6.88% for A*26:01, while the tumor sample showed LOH over *HLA-A*, with a methylation proportion of 25.19% for A*23:01 over the non-contaminated sample. When assessing across all titrations we observe a decrease in the methylation proportion of tumor A*23:01 allele following the expectation based on the tumor content., while the second allele that appeared from the 70% titration showed a methylation proportion close to the second allele of the normal sample (**Supplementary table 7**). In contrast, analyzing only the methylation proportion of the *HLA-A* gene as a whole only shows a decrease in methylation with decrease in tumor content. The additional information, coupled with the read imbalance (proportion of reads that cover each HLA allele, details below) helps us better analyze methylation differences in contaminated tumor samples.

Further, we observed clear errors from the HLA typing as we lowered the tumor purity. At 90% tumor purity, *HLA-B* typing changed from homozygous to heterozygous, with the second allele being distinct from the normal sample. Moreover, the re-alignment analysis shows an imbalance of 9:1 between both alleles, which resembles the tumor purity. Furthermore, at lower tumor purity, we could observe this imbalance decrease to 7:2 at 70% tumor purity, 5:2 at 50% and 2:1 at 30 and 10% tumor purity (**Supplementary table 7**). Taking this information into account, we called LoH at 90 and 70%, and flagged as suspicious at 50%. Finally, lower tumor purity also affected the number of cDMR to a ratio close to the tumor purity, further illustrating TumorLens’ ability to incorporate purity information.

Similar to methylation detection, variant detection is complicated by tumor purity at different concentrations. A somatic tumor variant occurring at 25% VAF is reduced at lower tumor concentrations and may eventually escape detection. For example at 70% purity the same variant would be reduced to 17.5% VAF (ie. ~5 reads in 30x), at 30% tumor content it is represented at only around 10% VAF (~3 reads in 30x). Similarly we observed a reduction of variant calls with a decrease of tumor purity from 50 to 100 SV for 70% to 10% tumor content, respectively. **Supplementary Table 6** shows the results for all variant classes. Similarly for SV we have to cope with CNV coverage changes that TumorLens automatically corrects for. For example a copy number (CN=3) would be reduced to 2.7 at 70% tumor content and down to 2 for 30% tumor content. This makes it harder to identify this event and report the ploidy correctly. Parallel to this change, the likelihood of false positive calls also increases at lower tumor purities. From 100 originally called CNV, TumoreLens was able to report 114 at 70% concentration but only 332 at 30% tumor concentration. We further observed a trend that CNV might be split into multiple events because of the decreasing tumor concentration and thus increase in noise levels of coverage.

Next, for LoH regions in the tumor samples, from 50% and lower tumor content, detection of LoH strongly decreased, in which we only detected a single LoH region in sample AOCS21 at 50% tumor content and none at 30 and 10%, even when previously detected nine chromosomes-wide LoH events in the same sample. Finally, we studied the effect of tumor content over HLA typing. Sample AOCS21 present single HLA allele in 5/8 typed *HLA* genes with one mistyped when compared to the normal, while sample AOCS9 present 4/8 typed *HLA* genes and one mistyped in both alleles (**Supplementary table 7**). The titration experiment showed that sample AOCS21 got two allele types “flipped to normal”, meaning agreeing with the normal sample and disagreeing with the tumor, at 70% tumor content, four flips at tumor content 50 and 30% and all alleles flipped at 10% tumor content. Notably, mistyping increased at tumor content 50%. For sample AOCS9, we detected two allele types “flipped to normal” starting at the 90% tumor content, one mistyping got corrected towards the normal sample and an additional mistyping occurred at both 90 and 70% tumor content. For 50% tumor content, most alleles show typing more similar to the normal than the tumor sample, thus showing that lower tumor content affects typing two-fold: 1) it flips the type towards the normal sample and 2) causes mistyping errors. To overcome this, we implemented a re-alignment approach (see **Methods** for details) that uses the normal sample typing as a reference and assesses the ratio between reported types to detect imbalances. **Supplementary Table 8** shows the alignment imbalance and how it correlates with decrease in tumor content.

Overall, we demonstrated the ability of the TumorLens pipeline to accurately recapitulate the genomic variants, differential methylation and typing, even at tumor purity as low as 50%. Thus, we were able to establish the accuracy of TumorLens to detect somatic mutations and methylation changes from long reads at different concentrations of tumor purities.

#### Lung cancer samples

We assessed large copy number variation (CNV) in the form of deletion and duplication. We observed that duplication is the most common CNV event among the samples, with seven samples having a duplication affecting the genes of interest. Furthermore, we detected somatic CNV events larger than 1Mb in 11 samples of the samples, and larger than 10Mb in eight samples.

In sample **lung_0,** a squamous cell carcinoma with 65% estimated tumor content, we detected duplications in Chromosome 5 (45Mb), 12 (21.8Mb, 11.8Mb), 14 (12.4Mb) and 15 (16.6Mb). One of the duplications in Chromosome 12 affects the six genes in the KLR locus (CN=3).

Then, for sample **lung_1**, also a squamous cell carcinoma (80% estimated tumor content), we detected a deletion of *TAP2,* which is a membrane-associated protein that encodes the protein antigen peptide transporter 2. These transporters are associated with antigen presentation and are thought to be responsible for conveying intracellular peptides into the endoplasmic reticulum for complex formation with class I MHC. Additionally, we detected a 50Mb duplication in Chromosome 7 that accounts for roughly 30% of the whole chromosome.

Sample **lung_2**, was removed from the analysis, as the quality control showed that these samples (tumor, normal or both) are likely contaminated.

Next, in sample **lung_4**, we detected a duplication on chromosome 6, affecting the *HSP90AB1* gene, which is frequently overexpressed in cancer cells ^2,3^. On other chromosomes, we also found a deletion affecting the *CTSB* gene, a known biomarker whose overexpression correlates with cancer invasion and metastasis ^4^ alongside a duplication of the *CD4* gene. Furthermore, we detected the most chromosomal aberrations among all analyzed samples with 24 large scale deletion and duplications (>10Mb) in Chromosomes 2, 3, 4, 7, 8, 9, 17, 20 and 22. Further, a duplication located in Chromosome 9 (9p21.3) (CN 3-4) affects the whole interferon gene cluster. It has been reported that copy number loss of the interferon gene cluster is associated with increased mortality and decreased overall survival in cancer ^5,6^, but not much has been reported for copy number gains. Additionally, the whole interferon gene locus also showed LoH on top of the duplication. We detected a total of 21 LoH regions greater than 10Mb across multiple chromosomes, affecting a plethora of immune-related genes including *CREB1, CTSB,* 13 Interferon genes, *HSPA5* and seven KIR genes from the APM gene list.

Finally, we performed allele-specific methylation analysis, focusing on the methylation differences of 74 immune-related genes involved in antigen processing and presentation (APM) and the human leukocyte antigen (HLA) system). We detected on average 5 genes showing allele specific methylation across the 13 lung cancer samples, with 25 of the 74 immune-related genes showing differential methylation in at least one comparison (**Supplementary table 11**) while 49 showing no methylation differences. Three samples showed over 10 genes showing allele- specific methylation (**lung_4**, lung_5, **lung_7**) while three samples showed one or none (**lung_1, lung_8, lung_A**), two of which have high tumor content, all observe low methylation differences genome-wide (**Supplementary Table 9**).

In sample **lung_4**, we detected the Cathepsin S gene (*CTSS*) as differentially methylated between the normal and tumor alleles, with the tumor alleles showing lower methylation (50% overall) contrasted to close to 90% in the normal. Elevated *CTSS* expression is observed in several cancers and is thought to contribute to tumor invasion and metastasis by promoting angiogenesis ^7^. Furthermore, also detected lower methylation in both tumor alleles of the Heat shock protein 90kDa alpha (*HSP90AA1*) with 30 and 22% methylation over the gene body and promoter in the tumor sample (allele 1 and 2 respectively) while close to 60% methylation the normal sample. *HSP90AA1* has been reported to be overexpressed in cancer cells and plays a role in the survival and proliferation of cancer cells and has been considered a potential target for cancer therapies ^8^.

Sample **lung_6**, an Adenocarcinoma with 80% tumor content showed 1,764 somatic SV, two somatic CNV and a very low proportion of cDMR, roughly 0.01%

Next, sample **lung_7** (80% tumor content) showed 14.77% of the genome differentially methylated, with most windows showing a lower methylation in the tumor sample compared to the normal. Among the affected genes, we found seven genes from the KIR locus. Further, we detected 12 large DEL including a 75Mb in Chromosome 3, a whole Chromosome 4 DEL, a 125 Mb DEL in Chromosome 5 and a 146 Mb DEL in Chromosome 6 and four large DUPs including a 96Mb duplication in Chromosome 8 (accounting for 65% of the chromosome). Among the affected genes, we found an over-representation of genes involved in antigen presentation (40 genes from APM and HLA locus)

Next, we compared the previous results to two squamous cell carcinoma samples (**lung_8** and **lung_A**) which each have 50% tumor content. The methylation analysis showed that only 97 (73 normal, 24 tumor) and 208 regions (199 normal, 9 tumor) showed differential methylation in samples **lung_8** and **lung_A** respectively, while no somatic genetic variant was detected in any of the samples. These results suggest that the tumor content of these samples is likely to be lower, resembling 10%-30% in the tumor content titration experiments. Notably, sample lung_0, a Squamous Cell Carcinoma with 65% tumor content, showed some methylation differences and somatic variants while sample **lung_9**, a Large Cell Carcinoma with 85% tumor content, showed 1991 somatic SVs, one somatic CNV and 1.48% of the genome classified as cDMR.

Finally in sample **lung_C** we detected allele-specific methylation within the Interferon Alpha 7 gene (*IFNA7*). One tumor allele exhibited reduced methylation (40% across the gene body and promoter), whereas the second tumor allele and both normal alleles exhibited ~80% or more methylation.

Across 12 samples, TumorLens identified differential methylation in approximately **8% of the genome** when comparing tumor and normal tissues, though five samples exhibited negligible differences (≤0.1% genome-wide). Furthermore, TumorLens detected loss of heterozygosity (LoH) in 4 of 13 samples, while large-scale duplications and deletions (>10 Mb) and large structural variants (SVs >50 kb) were each identified in 10 of 13 samples. Notably, every sample in the cohort displayed at least one large chromosomal copy number alteration (CNA). (**Supplementary Table 10**).

#### Tumor-only stomach samples

We utilized the ovarian sample AOCS21 to showcase the differences between tumor/normal and tumor-only analysis. For SV, we utilized STIX, a long-reads based annotation tool, to classify variants as pseudo-somatic if they were not present in the STIX database (see **Methods**). We identified 2,294 somatic SV on tumor/normal comparisons while the tumor-only analysis yielded 3,676 pseudo-somatic SVs. Comparison of the pseudo-somatic SVs to the somatic variants showed 340 SVs FP and not being somatic and 2,012 FN when taking the tumor/normal comparison and “truth”. Moreover, our result shows that the 2,012 SVs classified as somatic have been previously identified in individuals from the 1000 genomes project, while reporting only few FP.

For CNV and LoH, we utilized strict size filters to prioritize large events (> 1Mb). Using this strategy we identified all LoH somatic regions and 63% somatic CNVs (51 out of 81) in ovarian sample AOCS21.

In sample stomach_1, we identified three genes: PARM1, CAT, and CDKN2A, that exhibit significantly higher methylation compared to the normal panel. These genes are often reported as silenced or repressed in cancer. Specifically, PARM1 is frequently inactivated via mutations, deletions, or methylation; its loss disrupts cell cycle control and promotes unchecked proliferation, impacting prognosis and therapeutic response. CAT is known to be significantly downregulated in lung adenocarcinoma, where low expression is linked to poor prognosis. Finally, CDKN2A promoter methylation has been associated with tumor localization, histological subtypes, and Helicobacter pylori in gastric adenocarcinomas.

### Supplementary figures

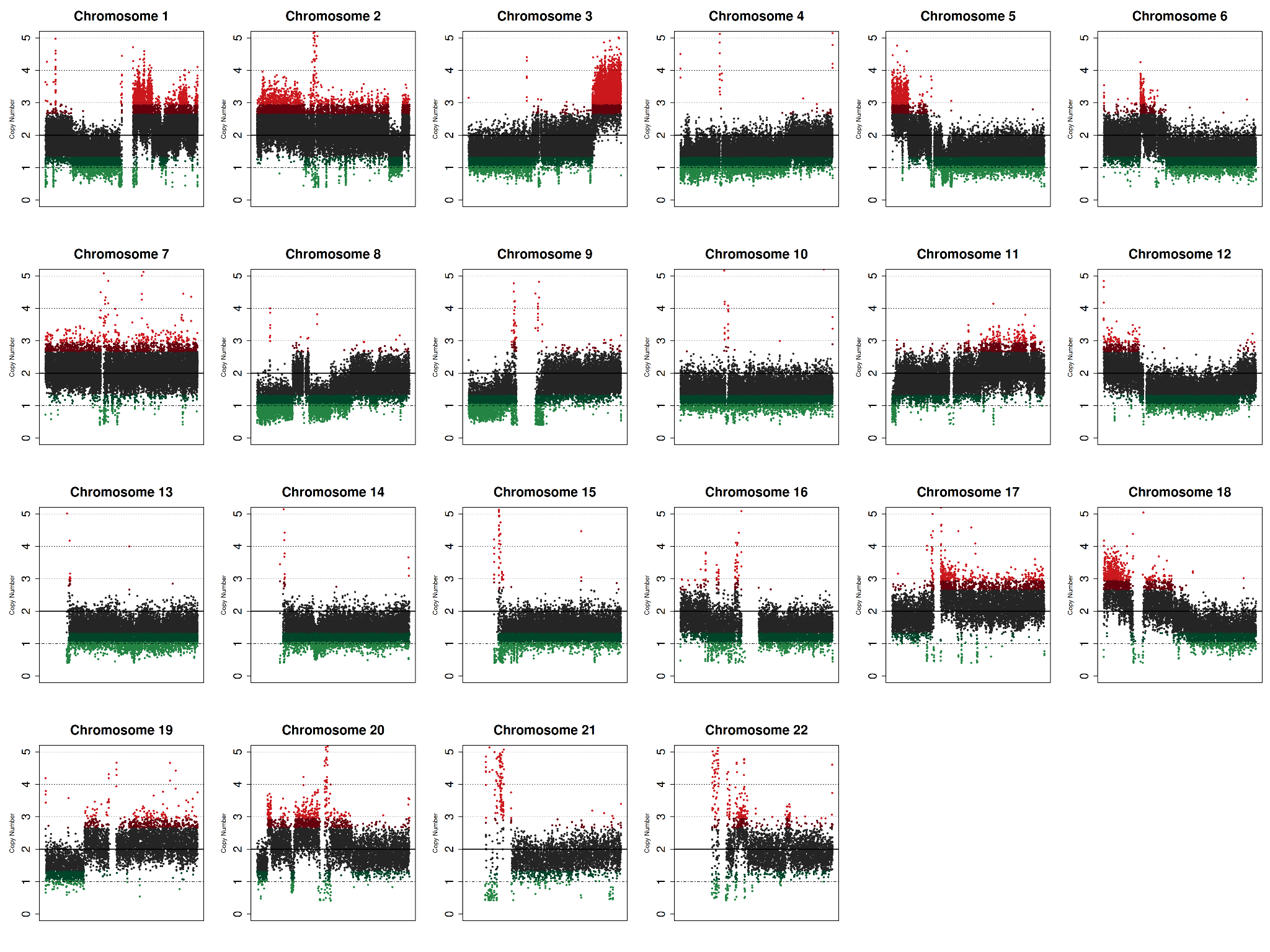

**Supplementary Figure 1.** Coverage plots of all the chromosomes in the tumor sample lung_5 (unspecified subtype, 70% tumor). The genomic regions depicted in green are below the threshold for deletion, while those in red are above the threshold for duplication.

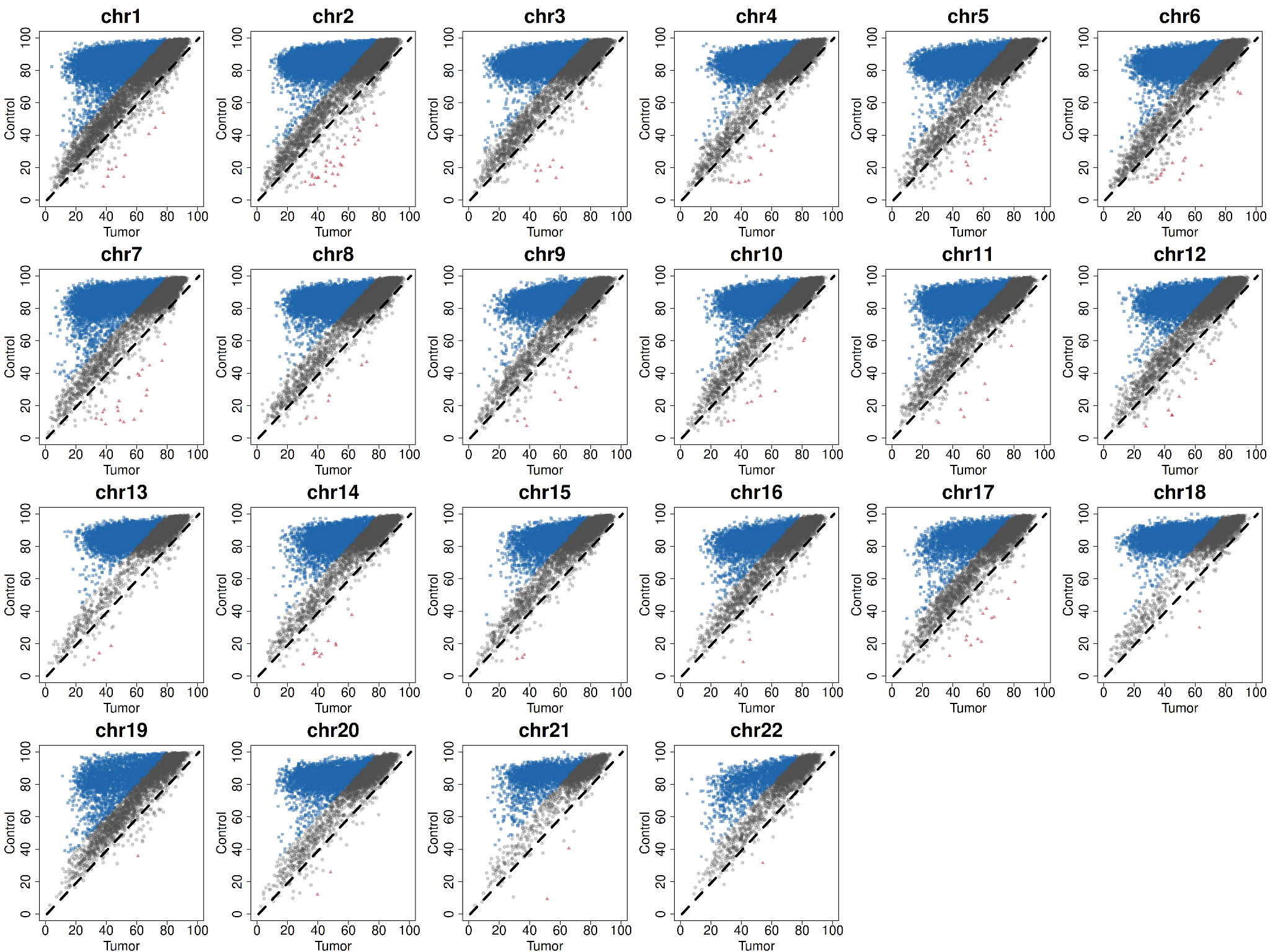

**Supplementary Figure 2.** Scatterplot comparing the proportion of methylation for the same genomic regions between tumor (x-axis) and normal (y-axis) in sample lung_5. The top-left corner represents those regions with high methylation in the normal and low in the tumor, while the bottom-right corner is *vice-versa.* In blue are genomic regions that have high methylation in the normal, showing over 33% points difference compared to the tumor. The red triangles represent regions where methylation is at least 33% higher in the tumor than in the normal . Both these regions are considered cancer differentially methylated regions (cDMRs)

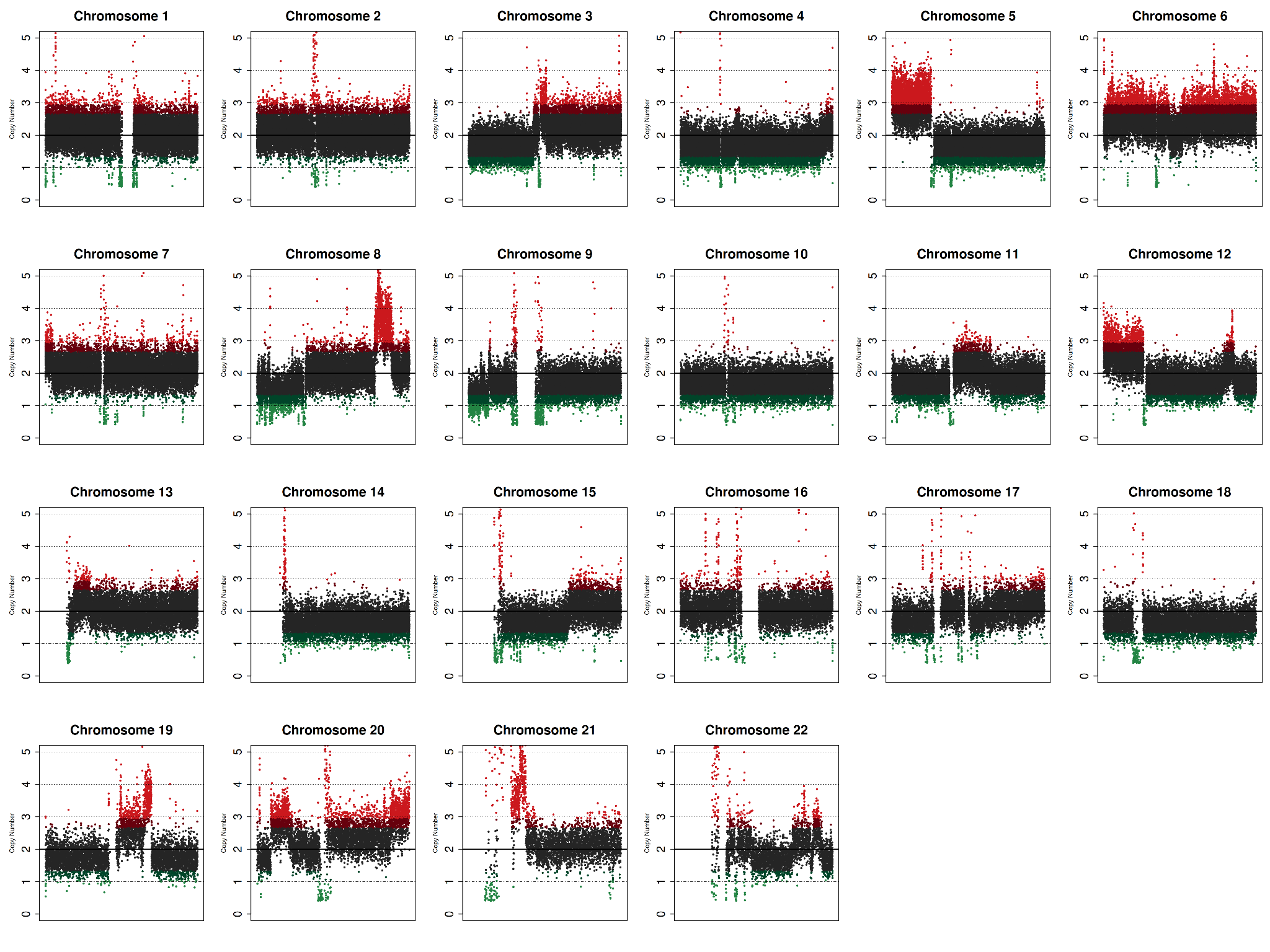

**Supplementary Figure 3.** Coverage plots of all the chromosomes in the tumor sample lung_B (large cell carcinoma, 65% tumor). The genomic regions depicted in green are below the threshold for deletion, while those in red are above the threshold for duplication.

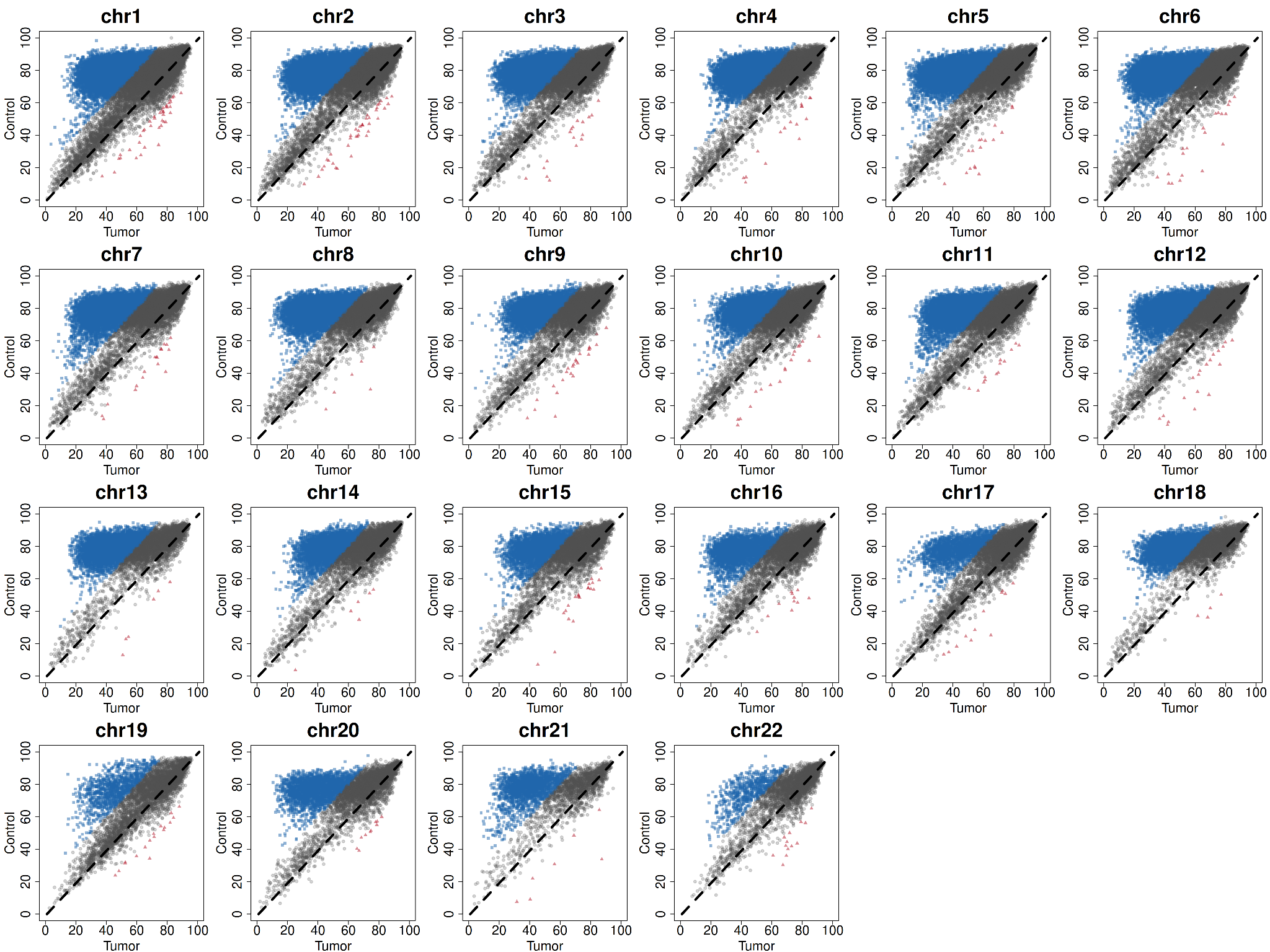

**Supplementary Figure 4.** Scatterplot comparing the proportion of methylation for the same genomic regions between tumor (x-axis) and normal (y-axis) in sample lung_B. The top-left corner represents those regions with high methylation in the normal and low in the tumor, while the bottom-right corner is *vice-versa.* In blue are genomic regions that have high methylation in the normal, showing over 33% points difference compared to the tumor. The red triangles represent regions where methylation is at least 33% higher in the tumor than in the normal. Both these regions are considered cancer differentially methylated regions (cDMRs)
